# Post-infectious inflammatory disease in MIS-C features elevated cytotoxicity signatures and autoreactivity that correlates with severity

**DOI:** 10.1101/2020.12.01.20241364

**Authors:** Anjali Ramaswamy, Nina N. Brodsky, Tomokazu S. Sumida, Michela Comi, Hiromitsu Asashima, Kenneth B. Hoehn, Ningshan Li, Yunqing Liu, Aagam Shah, Neal G. Ravindra, Jason Bishai, Alamzeb Khan, William Lau, Brian Sellers, Neha Bansal, Pamela Guerrerio, Avraham Unterman, Victoria Habet, Andrew J. Rice, Jason Catanzaro, Harsha Chandnani, Merrick Lopez, Naftali Kaminski, Charles S. Dela Cruz, John S. Tsang, Zuoheng Wang, Xiting Yan, Steven H. Kleinstein, David van Dijk, Richard W. Pierce, David A. Hafler, Carrie L. Lucas

**Author notes:** Equal contributors (Anjali Ramaswamy, Nina N. Brodsky, Tomokazu S. Sumida).

## Abstract

Multisystem inflammatory syndrome in children (MIS-C) is a life-threatening post-infectious complication occurring unpredictably weeks after mild or asymptomatic SARS-CoV2 infection in otherwise healthy children. Here, we define immune abnormalities in MIS-C compared to adult COVID-19 and pediatric/adult healthy controls using single-cell RNA sequencing, antigen receptor repertoire analysis, unbiased serum proteomics, and *in vitro* assays. Despite no evidence of active infection, we uncover elevated S100A-family alarmins in myeloid cells and marked enrichment of serum proteins that map to myeloid cells and pathways including cytokines, complement/coagulation, and fluid shear stress in MIS-C patients. Moreover, NK and CD8 T cell cytotoxicity genes are elevated, and plasmablasts harboring IgG1 and IgG3 are expanded. Consistently, we detect elevated binding of serum IgG from severe MIS-C patients to activated human cardiac microvascular endothelial cells in culture. Thus, we define immunopathology features of MIS-C with implications for predicting and managing this SARS-CoV2-induced critical illness in children.

## INTRODUCTION

Pediatric patients are largely spared of severe respiratory pathology associated with SARS-CoV2 infection; however, recent data has drawn attention to a severe and delayed post-SARS-CoV2 inflammatory response in children. This ‘multisystem inflammatory syndrome in children’ (MIS-C) presents in youth who had a mild or asymptomatic SARS-CoV2 infection roughly 4-6 weeks prior^1–9^. Symptoms in MIS-C patients vary and involve a systemic cytokine storm with fever, gastrointestinal, cardiac, vascular, hematologic, mucocutaneous, neurologic, and respiratory pathology, leading to critical illness with distributive/cardiogenic shock in up to 80% of patients and a 2% mortality rate^1^. Most patients with this syndrome are previously healthy with no co-morbidities and recover with supportive care and immune suppressive therapy. Further understanding the pathophysiology of this disease is imperative to predict, prevent, and optimally treat MIS-C in children exposed to SARS-CoV2.

Initial reports compared MIS-C with Kawasaki Disease (KD) because of the common presentation with fever, rash, and coronary aneurysms^2, 3, 5, 7, 8^. However, MIS-C predominantly affects older children with an increased prevalence among Black and Hispanic/Latino populations, whereas KD affects very young children with higher occurrence in East Asian populations. Moreover, MIS-C has distinct gastrointestinal symptoms, leukopenia, and high B-type natriuretic peptide, troponin, ferritin, and C-reactive protein, and it more frequently leads to shock^2, 10^. Acute MIS-C has been further characterized by high systemic inflammatory cytokines such as interleukin-1β (IL-1β), IL-6, IL-8, IL-10, IL-17, IFN-γ^<συπ>11</sup>^. Also reported is a cytokine profile indicative of NK, T cell, monocyte and neutrophil recruitment, mucosal immunity, and immune cell negative feedback^12^. Analysis of peripheral blood mononuclear cells (PBMCs) from MIS-C patients has revealed CD4, CD8, γδ T cell and B cell lymphopenia, with high HLA-DR expression on γδ and CCR7+ CD4 T cells, elevated CD64 expression on neutrophils and monocytes, and low HLA-DR and CD86 on monocytes and dendritic cells^11^. Neutralizing anti-SARS-CoV2 antibody responses in MIS-C closely resemble convalescent COVID-19, and recent studies also report higher complement C5b9 in serum and misshapen red blood cells, which are consistent with endothelial cell activation and clinical findings of distributive and cardiogenic shock^12–14^. Using panels of human antigens to screen for autoantibodies, acute MIS-C patients were described to have increased antibody binding to antigens associated with endothelium and heart development and other common autoimmunity targets as compared to healthy controls^12, 13^. As such, one of the dominant hypotheses to explain the immunopathology of MIS-C has been autoimmunity triggered by self-reactive antibodies produced in response to SARS-CoV2, as reported in KD where the presumed infectious trigger is often unknown^15–18^. This hypothesis, however, has not yet been directly tested.

Here, we report 15 cases of MIS-C and elucidate correlates of immunopathology using single-cell RNA sequencing with antigen receptor repertoire analysis, serum proteomics, and functional studies in a subset of acute and recovered MIS-C patients compared to healthy pediatric donors, adult COVID-19 patients, and healthy adults. We find innate and adaptive immune triggering during acute MIS-C that features elevated innate alarmins, acute inflammatory serum proteins, heightened cytotoxicity signatures, and expansion of IgG plasmablasts that correlate with serum antibody binding to cultured activated human cardiac microvascular endothelial cells in severe MIS-C.

## RESULTS

### Clinical characteristics distinguish moderate and severe MIS-C

Our clinical cohort includes 15 MIS-C patients, divided into severe and moderate groups based on clinical criteria (**Table S1**). Severe patients were critically ill, with cardiac and/or pulmonary failure (requiring vasoactive medication and/or significant respiratory support with positive pressure or mechanical ventilation), although not due to primary hypoxia. Most patients presented to care 4-6 weeks after peak adult COVID-19 hospitalizations (**Figure 1a**). Although a minority (6/15) of patients tested positive for SARS-CoV2 virus near the limit of detection during hospitalization, all of the patients had positive SARS-CoV2 serology. A majority of subjects presented with fever, gastrointestinal symptoms, and rash (**Figure S1a**). Two patients developed coronary aneurysms (**Figure S1a**), and most of the severe patients had depressed left ventricular heart function. All children received steroids, with a majority also receiving intravenous immunoglobulin (IVIG), and aspirin. Most severe patients also received vasoactive medications (epinephrine, norepinephrine, vasopressin, dopamine, and/or milrinone) and anakinra, an IL-1 receptor antagonist, along with heparin (enoxaparin) for anticoagulation and a course of antibiotics prior to negative culture results (**Figure 1b)**. Notably, PCA of clinical lab values separated severe and moderate MIS-C patients (**Figures 1c** and **S1b-c**). In keeping with prior studies, clinical labs for MIS-C patients showed high ferritin, b-type natriuretic peptide (BNP), troponin, c-reactive protein (CRP), soluble CD25, IL-6, and IL-10 (**Figure 1d** and **Table S2**). Severe patients also had high lactate, aspartate/alanine aminotransferases (AST/ALT), and creatinine, signifying the multi-organ involvement and shock state of their presentation (**Figures 1d**, **S1b** and **Table S2**). One critically ill patient (P1.1) had a throat culture that was positive for group A streptococcus (**Table S3**). All of the patients improved significantly, and all but one patient have been discharged home after an average of 7 days in the hospital.

**Figure 1.**
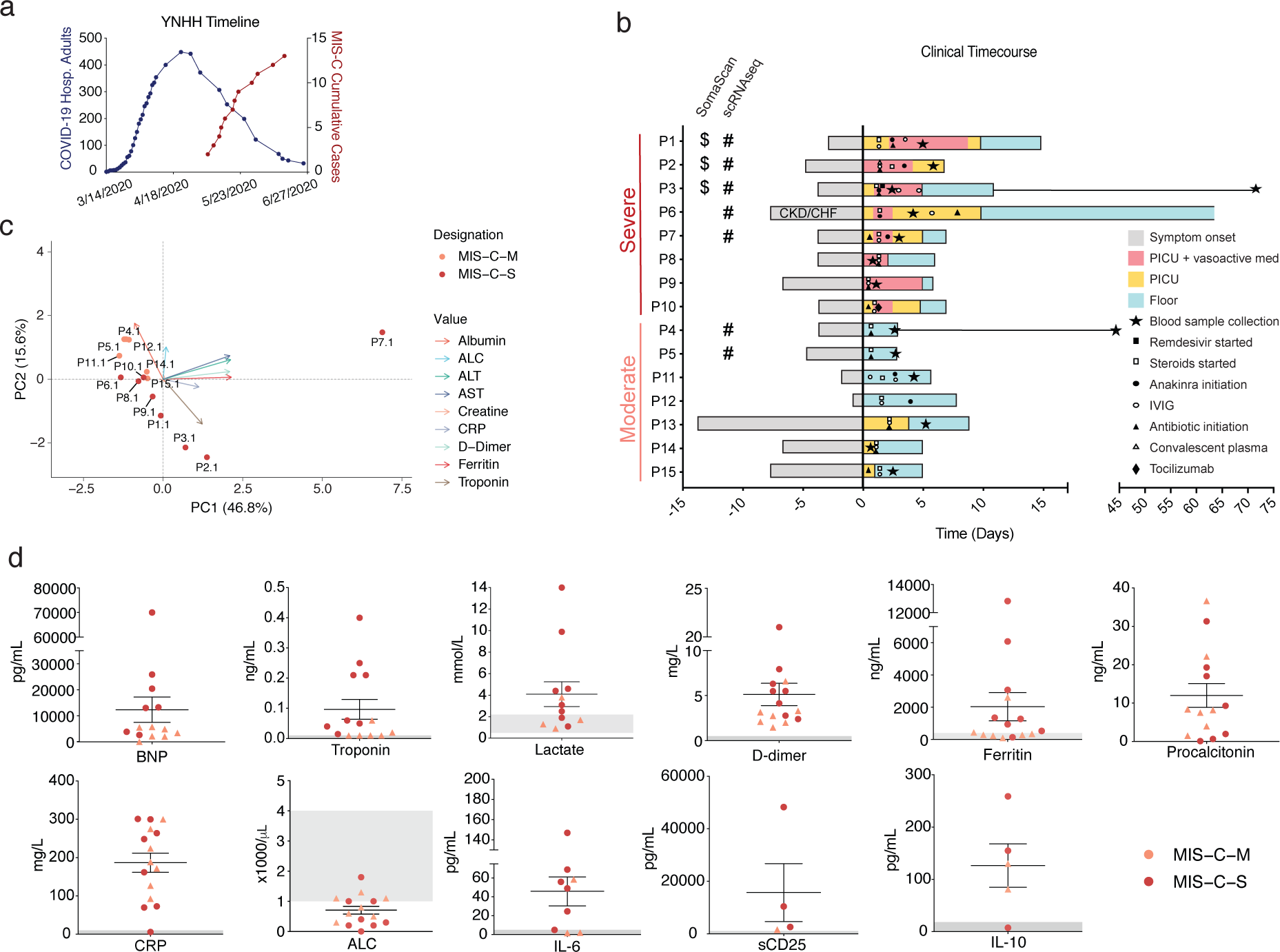
Clinical features of moderate and severe MIS-C. (**a**) Yale New Haven Hospital (YNHH) timeline of total daily adult COVID-19 hospitalizations (blue) and MIS-C cumulative cases (red). (**b**) Clinical time course of moderate and severe patients showing symptom onset and treatments relative to hospital admission (Day 0). (**c**) PCA biplot for clinical parameters, where available for MIS-C patients. Troponin was not measured for P13.1 was unavailable, and so this patient was excluded from PCA. (**d**) Clinical laboratory data for the indicated analyte. Normal range represented by gray shading. BNP: B-type natriuretic peptide; CRP: C-reactive protein; ALC: absolute lymphocyte count; AST: aspartate aminotransferase; ALT: alanine aminotransferase; WBC: white blood cells; CKD: chronic kidney disease; CHF: chronic heart failure.

### Altered MIS-C immune cell subsets with no evidence of active viral or bacterial infection

We surveyed the peripheral blood immune cell landscape of MIS-C by performing single-cell RNA sequencing (scRNAseq) on samples from six pediatric/child healthy donors (C.HD), seven MIS-C patients, and two recovered patients (MIS-C-R). We also incorporated samples from thirteen adult healthy donors (A.HD) and adult COVID-19 patients from early (COVID19-A: median of 7 days after symptom onset; n = 4) and late timepoints (COVID19-B: median of 16 days after symptom onset; n = 6) from our recent study (**Figure S1d** and **Table S4**)^19^.

We performed integrative analysis to harmonize all 38 single-cell gene expression (GEX) datasets, followed by graph-based clustering and non-linear dimensionality reduction using uniform manifold approximation and projection (UMAP) to visualize communities of similar cells. We resolved 30 distinct PBMC cell types (**Figure 2a-b and S2a-b**). Additionally, we performed Cellular Indexing of Transcriptomes and Epitopes by Sequencing (CITE-seq) on fresh PBMCs isolated from two MIS-C patients and three A.HD, allowing 189 surface antibody phenotypes to be resolved at a single-cell level together with GEX (**Table S5**)^20^. To annotate memory and naïve T cell GEX-based clusters, we exploited ADT signals for CD45RO and CD45RA. We also confirmed annotations of low-density neutrophils (retained after PBMC isolation) and mature NK cells using CD66d and CD57 markers, respectively (**Figure S2c**). We subsequently determined differences in cell type percentages among the pediatric cohorts **(Figures 2c** and **S2d).** Of note, naïve CD4 T cells were decreased in the peripheral blood of MIS-C patients compared to C.HD. Naïve B cells and platelets were markedly increased, and conventional dendritic cells (cDCs) and plasmacytoid dendritic cells (pDCs) were decreased in MIS-C compared to C.HD. We additionally leveraged scRNAseq GEX data to map significant changes in ligand-receptor connectivity in MIS-C compared to C.HD **(Figure S2e)**, finding that ligands and receptors involved in diapedesis and inflammation are coordinately up in MIS-C, including SELPG-ITGAM and MMP9-ITGB2.

**Figure 2.**
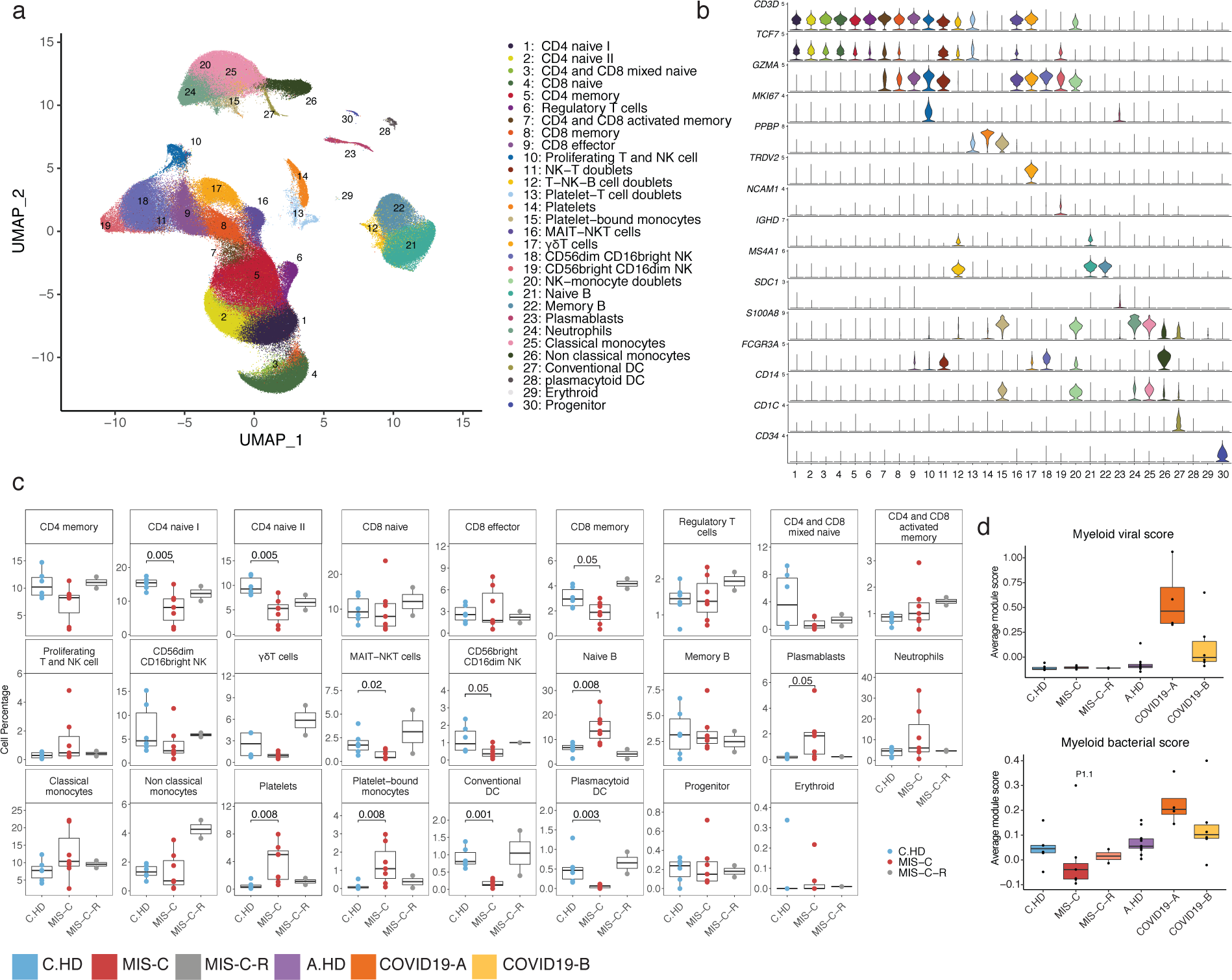
Altered MIS-C immune cell subsets with no evidence of active viral or bacterial infection. (**a**) Peripheral blood mononuclear cell (PBMC) UMAP of integrated samples from pediatric healthy donors, adult healthy donors, MIS-C patients, and COVID-19 patients. (**b**) Violin plots of key PBMC cell lineage markers. Y-axis represents normalized feature counts. (**c**) Distributions of peripheral blood cell frequencies across pediatric cohorts, based on cell types inferred from scRNA-seq. A non-parametric two-sided Wilcoxon test was used to assess statistical significance between the C.HD and MIS-C groups. (**d**) Donor distributions of viral and bacterial scores in myeloid cells. Module scores are calculated for each cell and averaged per donor.

To understand the possible viral or bacterial triggers for acute MIS-C onset, we evaluated well-defined signatures of respiratory viral and bacterial infections^21^. We detected a robust anti-viral signature in myeloid cells in COVID19-A but not in the MIS-C cohort (**Figures 2d** and **S2f**). Similarly, there was no evident bacterial signature in MIS-C compared to C.HD, with the exception of P1.1, who was confirmed to have strep throat at the time of MIS-C diagnosis (**Figure 2d** and **Table S3**). To determine whether an active herpesvirus could be found in patients with MIS-C, we created viral reference transcriptomes for Epstein Barr Virus (EBV) and Cytomegalovirus (CMV) for alignment of sequencing reads from our pediatric cohort. We did not identify counts aligning to these transcriptomes, with the few cells that appear positive for counts likely the result of mis-alignment (**Figure S2g**). Thus, peripheral blood cells in MIS-C patients show significant alterations, despite no direct evidence for an active viral or bacterial infection during acute illness.

### Elevated alarmin expression in myeloid cells with post-inflammatory phenotypic changes

To investigate innate immune contributions to MIS-C, we sub-clustered monocytes, neutrophils, and dendritic cells (**Figure 3a-b**). We additionally evaluated differentially expressed genes (**Table S6**) in neutrophils and monocytes, and found a shared up-regulation of the alarmin-related S100A genes^22^, in particular, *S100A8*, *S100A9*, and *S100A12* (**Figures 3c-d** and **S3d**). Consistent with previous scRNAseq studies, COVID19-A samples also showed an elevated S100A score compared to A.HD (**Figure 3d**)^19^. Pathway enrichment on shared down-regulated genes in monocytes and neutrophils revealed a significant reduction in antigen-presentation and processing, and as in COVID-19^23^, MIS-C patients had significantly reduced HLA class II (HLA-DP, DQ, and DR) expression (**Figure 3e and S3b-d,f)**. Moreover, MIS-C but not COVID-19 patients exhibited down-regulation of *CD86* (**Figures 3f** and **S3e**). This was also evident by flow cytometry of pediatric PBMCs (**Figure S3g**). To comprehensively define the serum proteome landscape in MIS-C, we profiled nearly 5,000 serum proteins in three MIS-C patients (P1.1, P2.1, P3.1) and four pediatric healthy donors using SomaScan technology (**Figure 3g, S3h**). Overall, there was a significant enrichment in myeloid-derived proteins among the differentially expressed proteins in serum (p = 1.7x10^-12^) (**Figure 3g**). Moreover, pathway analysis of differential proteins in the serum revealed enrichment of terms associated with ‘Cytokine-cytokine receptor interaction’, ‘Fluid shear stress and atherosclerosis’, and ‘Complement and coagulation cascades’, which are consistent with the inflammatory phenotype in the patients **(Figure 3h)**. To understand the impact of up-regulated serum proteins on immune cells, we performed connectivity analysis to link upregulated serum ligands with receptors expressed in PBMCs of MIS-C patients (**Figure S3i**). This analysis highlights CXCL10-CXCR3, which is known to be involved in leukocyte trafficking to inflamed tissues, as a potentially relevant axis in MIS-C^24, 25^. Thus, gene expression programs in myeloid cells from MIS-C patients are characterized by increased S100A alarmin expression and decreased antigen presentation that we hypothesize to be a post-inflammatory feedback response. Moreover, the serum proteome in MIS-C patients is consistent with inflammatory myeloid responses and potential endothelial cell activation.

**Figure 3.**
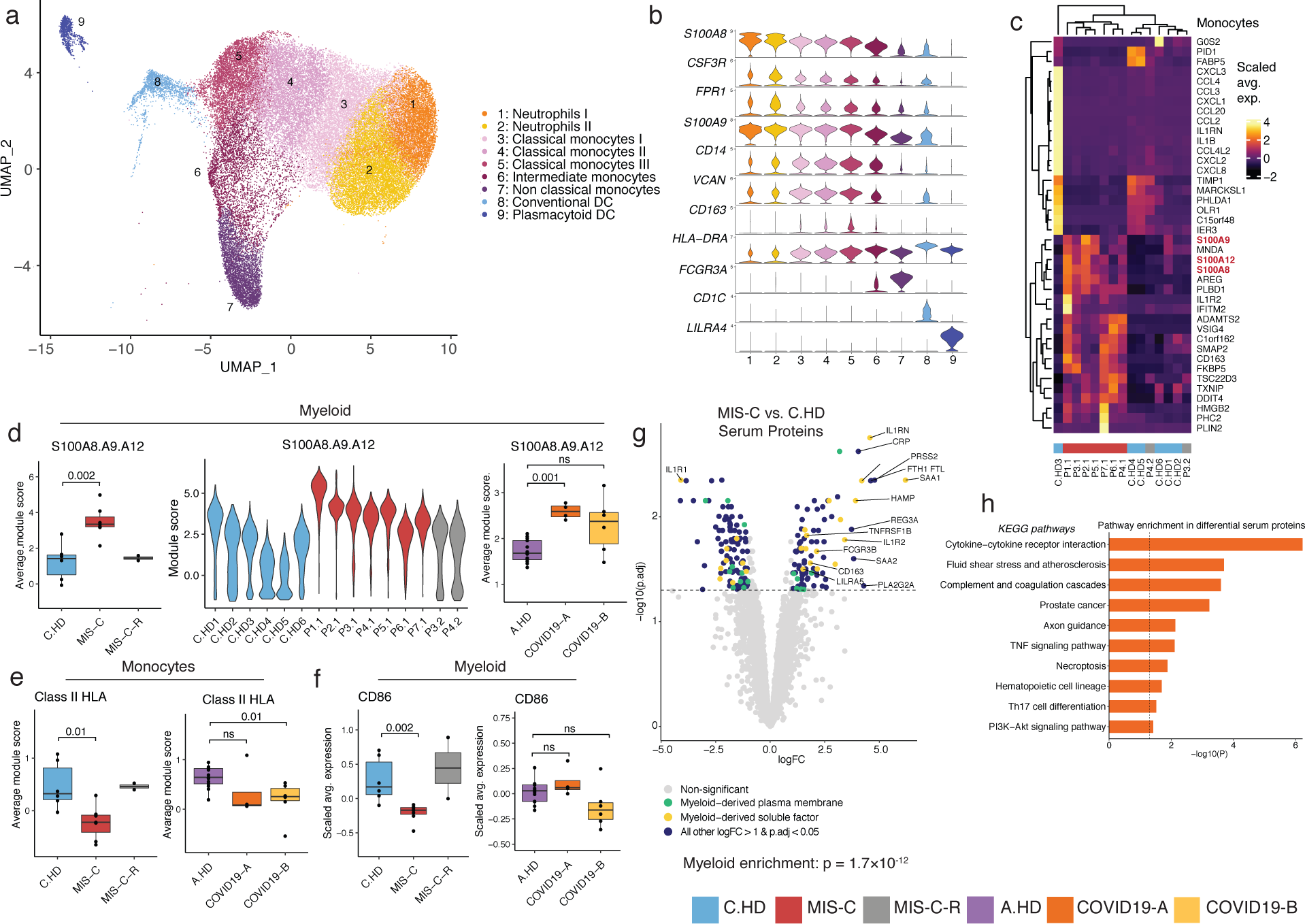
Innate inflammation in MIS-C with elevated myeloid alarmins in the S100A family. (**a**) Myeloid cell sub-clustering UMAP. (**b**) Key markers delineating myeloid clusters. (**c**) Heatmap representing top 20 up-and down-regulated differentially expressed genes in monocytes between MIS-C and C.HD. (**d**) A module score for *S100A8, S100A9,* and *S100A12* is computed across pediatric donors and adult healthy donors in all myeloid cells depicted in UMAP. As above, module scores are averaged per donor, and statistical significance between cohorts is computed using a two-sided non-parametric Wilcoxon test. (**e**) HLA class II score including *HLA-DP, DQ,* and *DR* molecules (see methods) is computed across adult and pediatric donors as above. (**f**) *CD86* expression depicted across pediatric and adult donors in myeloid cells. (**g**) Volcano plot showing differentially up-and down-regulated serum proteins between MIS-C (n=3) and pediatric healthy donors (n = 4). Molecule annotations are color-coded and genes of interest are labeled in black text. IL-1RN, an up-regulated protein in MIS-C, likely corresponds to anakinra treatment. Significance of enrichment is calculated using Fisher’s exact test. (**h**) Pathway analysis of differential proteins in serum analysis between MIS-C (n=3) and C.HD (n=4).

### Increased cytotoxicity genes in NK cells from MIS-C patients

To further define the T and NK cell states in MIS-C, we sub-clustered T and NK cells (**Figures 4a-b** and **S4a-b**). Marked differences were observed in cell type proportions of regulatory T cells and proliferating T and NK cells in MIS-C compared to C.HD (**Figure 4c**). To assess the potential for a superantigen response among T cells, we scored a defined signature of superantigen genes, which was not altered in MIS-C compared to C.HD (**Figure S4c)**^26^. However, differential gene expression analysis in the NK cell subset revealed a significant up-regulation of *PRF1*, *GZMA, and GZMH* cytotoxicity-related genes in MIS-C compared to C.HD, with high expression levels retained in recovered patients (**Figures 4d-e** and **S4d-e,g**). *CCL4*, produced by activated NK cells, was also up-regulated in MIS-C (**Figure S4f**). An analysis of T cells revealed a trend toward increases cytotoxicity gene expression in memory CD8 cells (**Figure S4d,h**). Additionally, *ITGB7*, an integrin subunit supporting lymphocyte infiltration of the gut through MADCAM1 binding^27^, was up-regulated in memory CD8 T cells (**Figure S4h**). Upon performing TCR diversity analysis, we found that a subset of MIS-C patients exhibited a decrease in memory and proliferating CD4 T cell clonal diversity, possibly indicating clonal expansion. However, no evidence of clonal expansion was seen in MIS-C CD8 cells. By contrast, COVID-19 patients exhibited markedly lower TCR diversity in both CD8 and CD4 cells (**Figure S4i**). Thus, NK cells, and to a lesser extent CD8 T cells, exhibit elevated cytotoxicity features with potential relevance for tissue damage.

**Figure 4.**
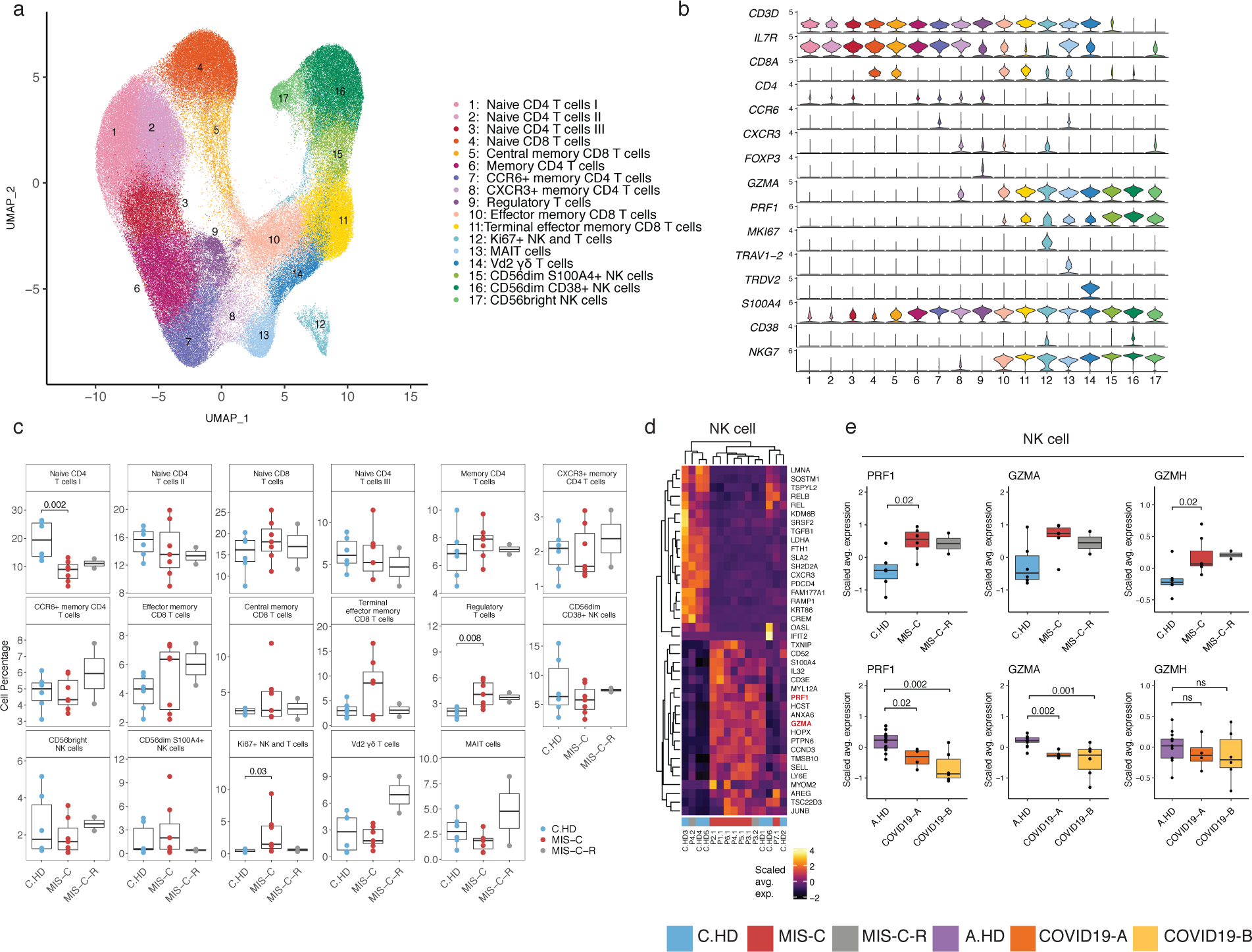
Increased cytotoxicity signatures in NK cells from MIS-C patients. (**a**) T cell sub-clustering UMAP. (**b**) Violin plot depicting key T and NK cell markers for cluster delineation. (**c**) T and NK compositions across pediatric cohorts. (**d**) Heatmap representing top 20 up- and down-regulated differentially expressed genes in NK cells between MIS-C and C.HD. Highlighted are genes associated with cytotoxicity. (**e**) *PRF1*, *GZMA, and GZMH* expression in NK cells in MIS-C compared to C.HD and MIS-C-R donors. Scaled average expression was calculated for each donor. A two-sided Wilcoxon test was calculated for statistical significance between cohorts.

### Elevated IgG plasmablasts in MIS-C

To investigate whether an ongoing humoral response could underpin acute MIS-C immunopathology, we sub-clustered annotated B cells (**Figure 5a-b**). We found a notable increase in proliferating (Ki67+) plasmablasts, which express apoptosis genes consistent with short-lived plasmablasts, in MIS-C compared to C.HD (**Figures 5c** and **S5a-b**)^28^. We performed differential expression analysis between naïve B cells of MIS-C and C.HD, and found an enrichment of the KEGG B cell signaling pathway among differentially up-regulated genes (**Figure S5c-d**). We next assessed antibody isotype, clonotypic diversity, and somatic mutation (SHM) of B cell receptors (BCRs). In memory B cells, the proportion of IgM B cells was increased in MIS-C (**Figure S5e**). Of note, the proportion of plasmablasts expressing IgG1 or IgG3 was elevated in MIS-C (**Figures 5d, S5f**), and a smaller proportion of plasmablast IgG clones in MIS-C and COVID-19 patients harbored mutated BCR variable regions (defined as >1% nucleotides mutated relative to germline) compared to age-matched controls (**Figure 5e**)^13, 29^. Moreover, a subset of MIS-C patients exhibits lower BCR clonal diversity – consistent with clonal expansion – when compared to C.HD, though this relationship is not as consistent as that between COVID-19 patients and A.HD (**Figure 5f)**.

**Figure 5.**
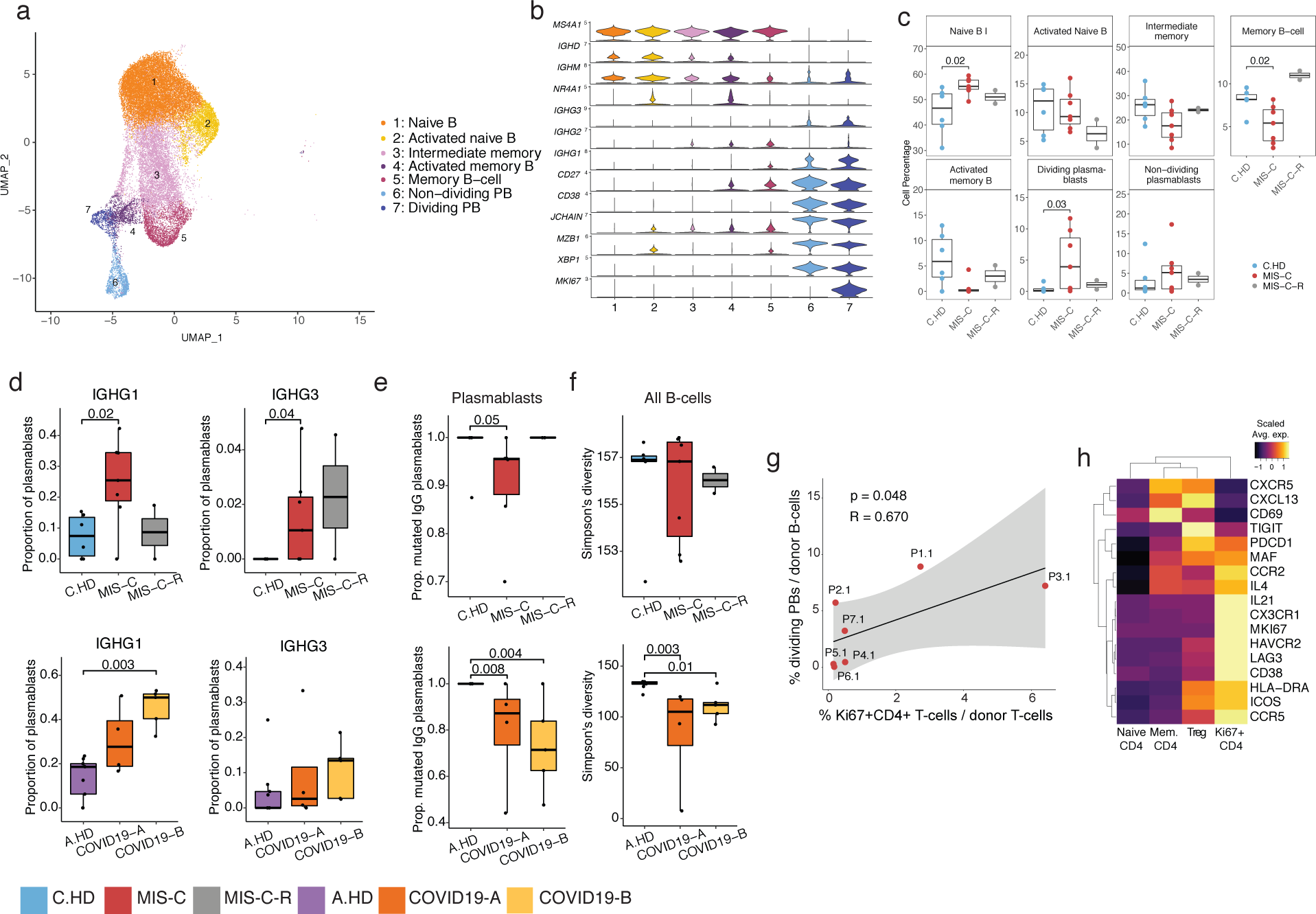
MIS-C patients have increased proliferating plasmablasts harboring IgG1 and IgG3 and a coordinated CD4 T cell response. (**a**) B cell sub-clustering UMAP. (**b**) Violin plots for key B cell markers delineating naïve, memory, and plasmablast subsets. (**c**) Distributions of B cell frequencies within total B cells across donors. (**d**) *IGHG1* and *IGHG3* isotype frequencies as a proportion of plasmablasts (dividing and non-dividing) are depicted across donors. Two-sided Wilcoxon rank sum tests were used to calculate significance. (**e**) Proportion mutated *IGHG* clones in plasmablasts. (**f**) Simpson’s diversity in all B cells computed across cohorts in pediatric cohorts (top) and adult cohorts (bottom). Significance calculated as above. (**g**) Correlation of percentage dividing plasmablasts/total B cells versus percentage Ki67+ CD4 cells/total T cells within the MIS-C cohort. Ki67+ CD4 cells defined as CD4+ cells within the Ki67+ NK and T cell cluster (see Figure 4a). Linear regression is shown with 95% confidence interval (gray area). Correlation statistics by two-tailed Spearman rank correlation test. (**h**) Heatmap showing differential gene expression across four subsets of CD4+ T cells, including samples from all 38 human subjects.

To examine potential drivers of this plasmablast response, we looked at correlates in the CD4 T cell response. Indeed, we found a positive correlation between proliferating Ki67+ CD4 T cells and Ki67+ plasmablasts (**Figures 5g** and **S5g**). Gene expression analysis of Ki67+ CD4 T cells revealed low CXCR5, but high *ICOS*, *PDCD1*, *MAF*, and *IL21* as well as chemokine receptors for homing to inflamed tissue including *CCR2*, *CX3CR1*, and *CCR5* (**Figure 5h, S5h)**. These cells appear to be phenotypically similar to T peripheral helper cells seen in some autoimmune conditions^30^. Together, these data indicate that plasmablasts in MIS-C patients are expanded, correlate with proliferating CD4 T cells with putative B cell-helper function, and more frequently harbor IgG1 and IgG3 antibody isotypes compared to C.HD.

### Evidence for immunopathology in severe MIS-C patients

Severe and moderate MIS-C patients are clinically distinct (**Table S1**). Thus, we hypothesized immunopathology features in severe patients (here called MIS-C-S) would be more pronounced than moderate (here called MIS-C-M) and stratified the subjects for further analysis.

To begin to assess TCR repertoire skewing from prior exposures, possibly in response to previous SARS-CoV2 infection, we assessed memory T cell compartments using PCA of *TRBV* gene usage. Both CD4 and CD8 memory T cells exhibited significant skewing of the V-beta repertoire, with TRBV11 significantly enriched in MIS-C-S (n=4) compared to C.HD (n=6) or MIS-C-M (n=2) in both compartments (**Figure 6a, S6a-b**). COVID19-A and COVID19-B memory T cells did not exhibit a separation from A.HD (**Figure S6c-d**), possibly indicating a specific skewing event in the memory compartment in MIS-C. Moreover, effector CD8 T cells exhibited a significant up-regulation of *PRF1* and increases in *GZMA* and *GZMH* when comparing MIS-C-S with C.HD (**Figures 6b**). *LAG3*, an inhibitory receptor up-regulated upon T cell activation, was also increased in effector CD8 T cells (**Figure S6e**). Based on data in **Figure 5g** demonstrating a coordinated CD4 T cell and plasmablast response in some MIS-C patients, we assessed correlations further in MIS-C-S. Indeed, we found that patients with low B cell clonal diversity also had low combined Ki67+ and CD4 memory T cell diversity, suggestive of coordinated clonal expansion (**Figure 6c**). Furthermore, in MIS-C-S, we found increased plasmablast frequencies, decreased total B cell clonal diversity, and an increased proportion of mutated IgG clones, consistent with a more robust B cell response in these patients (**Figure 6d**). Consistent with these findings, the proportion of *IGHG1*- and *IGHG3*-plasmablasts was higher in MIS-C-S patients compared to controls (**Figure S6f**).

**Figure 6.**
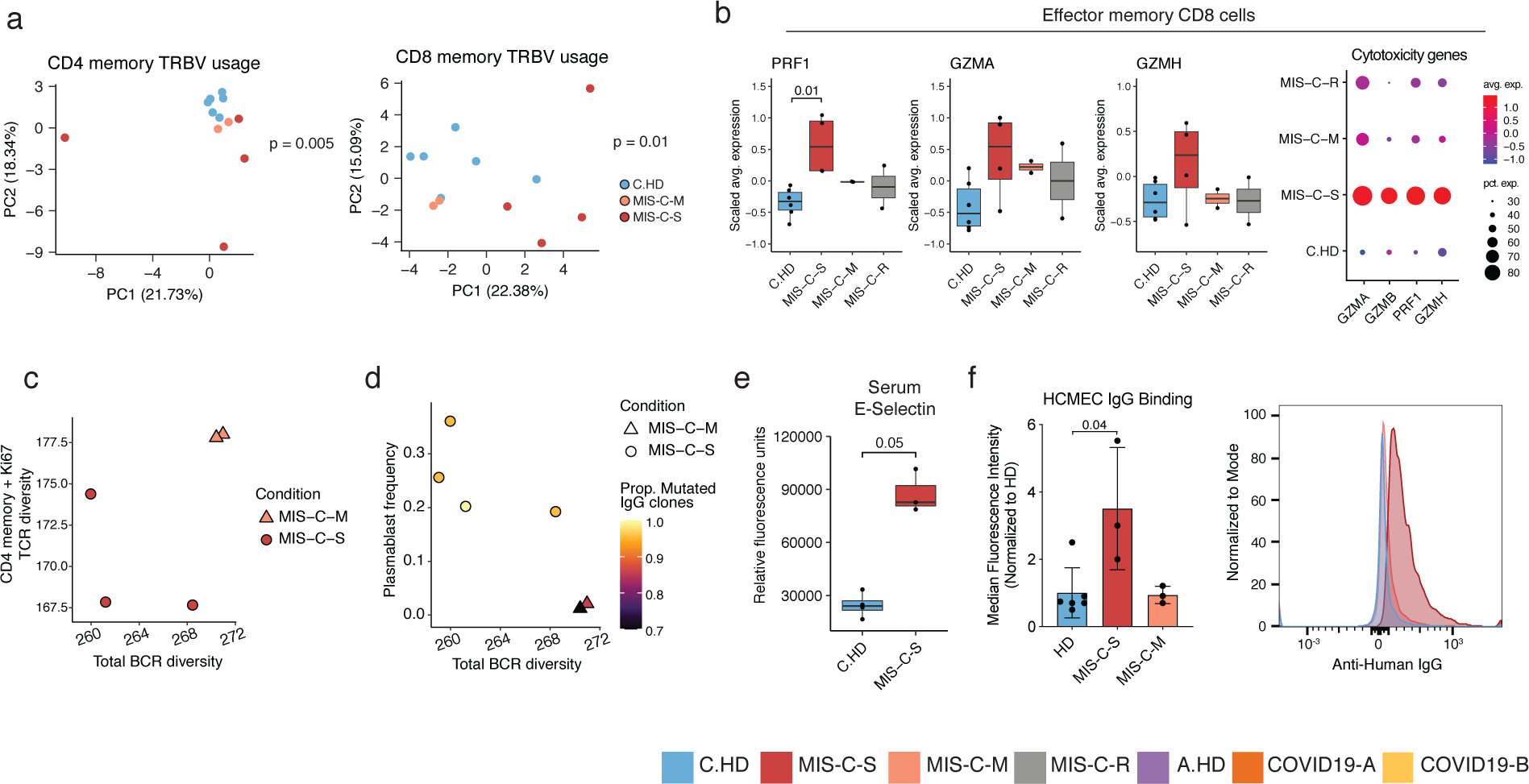
Distinct features of severe versus moderate MIS-C. (**a**) PCA of TRBV usage in CD4 and CD8 memory cells in the pediatric cohort. Statistical significance calculated by permutation test (see methods) (**b**) *PRF1* and *GZMA* expression in effector memory CD8+ T cells along with dot plot depicting relative average expression and percent expression for four cytotoxicity genes (right). Wilcoxon rank sum tests were used to calculate significance. (**c**) Correlation between BCR diversity and TCR diversity relating to combined Ki67+ and memory CD4 T cells. P7.1 was excluded from TCR analysis due to low cell numbers (see methods). (**d**) B-cell diversity, plasmablast frequency, and proportion of mutated *IGHG* within MIS-C cohort. (**e**) Serum E-selectin in pediatric healthy and MIS-C donors. (**f**) Mean fluorescence intensity (normalized to average HD) of serum IgG binding to cultured human cardiac microvascular endothelial cells (HCMEC) by flow cytometry (left). A non-parametric Wilcoxon rank sum test was used to calculate significance. MIS-C-S (n=3; P1-3); MIS-C-M (n=2; P4-5); and HD (n=5; 1 C.HD and 4 A.HD). Representative histogram on the right is from P3 prior to IVIG treatment (right).

Given the distributive and cardiogenic shock in MIS-C, we aimed to investigate endothelial cell involvement. Mining the serum proteomics data from **Figure 3g** further, we noted that endothelial E-selectin, a molecule known to be expressed on inflamed endothelial cells, was also markedly up-regulated in MIS-C-S serum (**Figure 6e**)^31^. Next, we assessed a possible autoantibody response directed at endothelial cells. We examined binding of MIS-C serum antibodies to cultured human cardiac microvascular endothelial cells (HCMEC). Indeed, IgG from severe (P1.1, P2.1, P3.1, the latter being pre-IVIG) but not moderate (P4.1, P5.1, P11.1) MIS-C patients bound activated endothelial cells (**Figure 6f**), consistent with a potential autoimmune process. Thus, T and B cell clonal expansion, as well as cytotoxic gene expression signatures in CD8 T cells, appear to correspond with severe MIS-C. Importantly, we provide functional evidence for MIS-C autoantibody binding to activated endothelial cells relevant for severe disease pathology.

## DISCUSSION

We describe our findings from comprehensive analysis of MIS-C patients using single-cell RNA sequencing, antigen receptor repertoire analysis, serum proteomics, and *in vitro* assays.

Separation of MIS-C into moderate and severe groups based on clinical criteria uncovered signals of disease pathogenesis that otherwise would not have emerged. As previously reported^11^, myeloid cells in MIS-C express lower HLA class II and *CD86*, molecules involved in antigen presentation, likely as a compensatory post-inflammatory feedback response. We additionally identified an up-regulation of alarmin genes including subunits of calprotectin (*S100A8* and *S100A9*) as well as EN-RAGE (*S100A12*) that function to amplify inflammatory responses. Moreover, unbiased surveying of nearly 5,000 serum proteins revealed differential abundance of many acute phase and myeloid-derived inflammatory proteins, as well as elevated endothelial E-selectin, consistent with systemic inflammation and endothelial activation.

What drives the cytokine storm and multi-organ damage in MIS-C? In addition to possible innate drivers described above, our analysis of lymphocytes from MIS-C patients points to three striking findings. First, NK cells and, to a lesser extent, CD8 T cells express elevated perforin, granzyme A, and granzyme H, cytotoxic molecules of relevance for tissue damage. In contrast to granzyme B, granzyme A is known to cleave pro-IL-1β and may directly contribute to inflammation beyond its cytotoxic function^32, 33^. Second, B cells have an expansion of proliferating plasmablasts that fits with a potential humoral response weeks after clearance of SARS-CoV2, raising the possibility that these are autoreactive expansions of antibody-secreting cells. Third, evaluation of severe MIS-C patients identified TCR repertoire skewing among memory T cells, evidence of clonal expansion and somatic hypermutation within B cell populations, and measurable binding of serum IgG to activated cardiac endothelial cells in culture. The plasmablasts expanded in MIS-C show evidence of being short-lived with upregulated pro-apoptotic genes, which may help explain the self-resolving nature of pathology. Collectively, our data support a model in which prior SARS-CoV2 infection causes lasting immune alterations that set the stage for development of an acute and life-threatening post-infectious inflammatory episode in a fraction of children and adolescents (**Figure S7**).

Three main possibilities exist to explain the rare occurrence of MIS-C. First, a rare genetic predisposition could underlie disease, and future genomics investigations will be revealing on this front. Second, similarly to rheumatic heart disease, the infectious trigger could elicit adaptive immune responses that, on rare occasion, cross-react with self-antigens. The rapid resolution of inflammation in MIS-C may go against this theory. Third, a rare combination of SARS-CoV2 infection followed by a second microbial trigger could drive the acute MIS-C inflammatory episode. We did not find evidence of herpesvirus reactivation or peripheral blood signatures of ongoing viral or bacterial infections. Nonetheless, a tissue-specific response to an infectious trigger remains plausible, and the common feature of abdominal pain early in the course of MIS-C is suggestive of potential gut involvement and consistent with elevated *ITGB7* in T cell subsets. Further work is required to define contributions of each of these potential triggers.

The determinants of whether a child with MIS-C develops moderate or severe disease are also unknown and may relate to prior SARS-CoV2 viral load and immune repertoire shaping and/or differences in the putative secondary MIS-C-triggering event. Although patients with severe disease have more potential autoantibody as measured by IgG binding to cultured endothelial cells, whether this is causative of severe disease or a result of increased tissue destruction and autoantigen exposure cannot currently be determined. Moreover, by necessity, MIS-C patients are promptly treated with anti-inflammatory drugs, complicating analysis of the disease state, though we have attempted to use existing knowledge on methylprednisolone effects to inform our findings (Figure S3f)^34^. Although our findings in MIS-C require larger patient numbers and further testing, ideally including animal modeling, they have important implications for predicting, preventing, and treating MIS-C and offer potential paths forward for diagnostic and prognostic testing. With new waves of SARS-CoV2 outbreaks on the horizon and eventual vaccination against SARS-CoV2 in children as a critical goal, a better understanding of MIS-C drivers and immunopathology is urgently needed. Our data implicate innate and adaptive immune triggering with direct relevance for tissue destruction during acute MIS-C.

## Data Availability

Raw and processed data pertaining to pediatric and select adult samples (A.HD1-3) in this paper are available at GEO: GSE166489 and FASTgenomics. Data pertaining to other samples are available from previous studies (Pappalardo et al., Science Immunology 2020, Unterman et al., medRxiv 2020). Code is made available in https://github.com/LucasiteLab/MIS-C_scRNAseq and FASTgenomics.

## ACKNOWLEDGMENTS

The authors thank the patients and their families for participation. We are also grateful to the physicians, nurses, and hospital staff who helped care for the patients and obtain samples. The authors thank J. Pober and D. Jane-Wit for critical input, R. Montgomery for support, and G. Wang and C. Castaldi at Yale Center for Genome Analysis for 10x Genomics library preparation and sequencing services; K. Raddassi for processing scRNA-seq samples; J.L. Pappalardo for providing us the healthy adult scRNA-seq data; R. Sparks and L. Failla for assistance providing healthy pediatric samples for SomaLogic. We also thank Ms. M. Bucklin and D. Murdock for feedback and discussions. This research was supported by grants to C.L.L. from NIAID 3R21AI144315-01A1S1 and Yale University. R.W.P is supported by NHLBI 1K08HL136898-01A1. AR was supported by NIAID 5T32AI007019 and NSF Graduate Research Fellowship.

## AUTHOR CONTRIBUTIONS

A.R., N.N.B., and C.L.L. conceptualized the study. N.N.B., V.H., A.J.R., R.S., and J.C. consented patients and healthy donors. N.N.B, V.H., H.C. and M.L. collected blood samples and clinical information from all patients. N.N.B. and A.J.R. performed serum, plasma and PBMC isolation and cryopreservation. T.S.S., and H.A. perform scRNA-seq and CITE-seq sample preparation and cDNA generation; A.R., T.S.S., M.C., H.A., A.U., A.S., S.K., D.V.D., and C.L.L analyzed scRNA-seq gene expression data and CITE-seq data with the help of N.K. and D.A.H.; K.H. and S.K. analyzed single cell BCR data; N.L. and X.Y. analyzed single cell TCR data; W.L., B.S., N.B., J.C. and J.T. analyzed somascan serum proteome data; M.C. performed flow cytometry analysis for patients’ PBMCs; N.N.B., A.K., and R.P. performed endothelial cell experiments; A.R., N.N.B., and C.L.L. wrote the manuscript with input from all authors; C.L.L. supervised the overall study.

## METHODS

### Human subjects

All human subjects in this study provided informed consent in accordance with Helsinki principles for enrollment in research protocols that were approved by the Institutional Review Board of Yale University. Patients were enrolled from Yale New Haven Children’s Hospital (New Haven, CT) and Loma Linda Children’s Hospital (Loma Linda, CA). Blood from healthy donors was obtained at Yale under approved protocols.

### COVID-19 samples

COVID19-A and COVID19-B samples were provided by Unterman et al. 2020^19^. COVID19-A and COVID19-B blood draws were taken post-hospitalization, with a median time elapsed between timepoints of 4 days. Two samples, A.COV5, and A.COV6, only correspond to timepoint B.

### Blood sample processing

Human PBMCs were isolated by Ficoll-Paque PLUS (GE Healthcare) or Lymphoprep (STEMCELL Technologies) density gradient centrifugation, washed twice in PBS, and resuspended at 10^6^ cells/ml in complete RPMI 1640 (cRPMI) medium (Lonza) containing 10% FBS, 2 mM glutamine, and 100 U/ml each of penicillin and streptomycin (Invitrogen). PBMCs were used fresh or cryopreserved in 10% DMSO in FBS and thawed prior to use. Serum was isolated by centrifugation of serum tubes and saving the supernatant in aliquots which were flash frozen in liquid nitrogen prior to cryopreservation in -80C.

### Single-cell RNA-sequencing

Cryopreserved PBMCs were thawed in a water bath at 37°C for ∼2 min without agitation, and removed from the water bath when a tiny ice crystal still remains. Cells were transferred to a 15 mL conical tube and the cryovial was rinsed with growth medium (10% FBS in DMEM) to recover leftover cells, and the rinse medium was added dropwise to the 15 mL conical tube while gently shaking the tube. Next, growth medium was added at a speed of 3-5 ml/sec, achieving a final volume of 13 mL.

Fresh or thawed PBMCs were centrifuged at 400 g for 8 minutes at RT, and the supernatant was removed without disrupting the cell pellet. The pellet was resuspended in 1X PBS with 0.04% BSA, and cells were filtered with a 30 μM cell strainer. Cellular concentration was adjusted to 1,000 cells/μl based on the cell count and cells were immediately loaded onto the 10x Chromium Next GEM Chip G, according to the manufacturer’s user guide (Chromium Next GEM SingleCell V(D)J Reagent Kits v1.1). We aimed to obtain a yield of ∼10,000 cells per lane.

For CITE-seq staining, lyophilized Total-seq C human cocktail (BioLegend) (**Table S2**) was resuspended with 35 μL of 2% FBS in PBS vortexed for 10 sec and incubated for 5 min at RT. To pellet the aggregated antibodies, rehydrated antibody cocktail was centrifuged at 20,000g for 10 min just before adding to the cells. PBMCs were resuspended with wash buffer at the concentration of 10-20 x 10^6^ cells/ml, and 0.5 x 10^6^ cells were used for further staining. Cells were incubated on ice for 10 mins with 5ul of Human Fc block and 5 μL of TrueStain Monocyte Blocker (Biolegend). Next, 10-20 μL (0.1-0.2 x 10^6^ cells) were aliquoted into a new tube and incubated on ice for 30 mins with 5 μL of Total-seq C antibody cocktail prepared as above. Cells were washed twice with wash buffer and third wash was with 2% FBS in PBS, then resuspended in 1X PBS with 0.04% BSA at 1,000 cells/ul and loaded onto the 10x Chromium Chip G, as described above. cDNA libraries for gene expression, CITE-seq, and TCR/BCR sequencing were generated according to manufacturer’s instructions (Chromium Next GEM SingleCell V(D)J Reagent Kits v1.1). Each library was then sequenced on an Illumina Novaseq 6000 platform. The sequencing data was processed using CellRanger v3.1.0.

### PBMC single-cell RNA sequencing analysis

Pediatric healthy donor, MIS-C, longitudinal recovered MIS-C, adult healthy donor, and adult COVID-19 PBMC CellRanger outputs were analyzed using the *Seurat* v3.2.1 package^35^. These data were filtered, log-normalized, integrated, and scaled prior to dimensionality reduction and cluster identification. For each dataset, we filtered out genes that were expressed in fewer than 5 cells, and we removed low quality cells which have over 10% mitochondrial gene content and contain fewer than 200 features. To remove batch-and single-donor effects, we integrated all 38 samples into one dataset using Seurat’s reference-based anchor finding and integration workflow, which is recommended by Seurat for integrating large numbers of datasets. We chose an adult healthy donor sample (A.HD3) and a MIS-C patient sample (P1.1) as references for anchor finding and integration, and used 2000 anchors and the first 30 principal components (PCs) for the integration steps. To reduce dependence of clustering on cell-cycle heterogeneity, we scored cells for cell-cycle phase based on a defined set of phase-specific genes and regressed out these genes during the scaling step.

Principal component analysis was performed on the scaled dataset. To define the number of principal components (PCs) to use we applied the elbow plot method and we also tested different numbers of PCs to evaluate the effects on the separation of distinct cell lineages. Based on these determinations, we chose the first 30 PCs for nearest neighbor identification and a clustering resolution of 1.0 for cluster finding. Finally, we chose UMAP as a non-linear dimensionality reduction approach to visualize clusters. To define clusters, we calculated differentially expressed genes specific to each cluster using Wilcoxon rank sum test. Cluster specific markers were found that had an absolute logFC of at least 0.25, an adjusted p-value of less than 0.05, and were expressed in a minimum of 25% of cells in either cluster being compared. Dead and dying cell clusters were identified as those with high mitochondrial gene content, low number of unique genes, and mitochondrial genes as the top cluster-specific differentially expressed genes. After removing cells belonging to these clusters, the data was re-processed using the same parameters above and clusters were annotated using cluster specific differential expression. All scRNA-seq analysis was done using R version 4.0.2^36^. All scRNA-seq plots were done using *ggplot2* v3.3.2^37^.

### CITE-seq analysis

Of the 38 samples, 5 included CITE-seq data. After integrating all of the datasets in both the PBMC and sub-clustering analyses, we used a subset of our Seurat object corresponding to these 5 donors, and overlaid ADT information onto the GEX-based UMAP for cluster validation. ADT data was log-normalized prior to plotting feature counts.

### Connectivity mapping

The Connectome v0.2.2 package was used to generate a network analysis of ligand-receptor interactions predicted to be up- or down-regulated in MIS-C compared to C.HD^38^. PBMC clusters were included that were represented in both MIS-C and C.HD groups. We excluded clusters containing doublets and clusters where the sum of cells was fewer than 75 cells in either MIS-C or C.HD. To minimize differences in connectivity due to cell compositions between cohorts, we down-sampled our dataset. Specifically, for each cluster, we computed the sum of cells belonging to MIS-C patients or C.HD, and used the minimum of the two values to randomly sample cells within MIS-C or C.HD in the relevant cluster.

To annotate ligands and receptors, we used a list of annotated human ligand-receptor pairs sourced from the FANTOM5 database appended with immunological ligands and receptors created in a recent scRNAseq study^19^. Connectomes were created for each down-sampled cohort. An edge is determined as a ligand-receptor pair that is expressed in respective clusters at a level greater than 5% of cells, and edge-weights are determined as the sum of the scaled expression values of the markers^38^. The two connectomes were then compared to create a fold-change connectome, and this was then filtered to only differentially expressed genes between MIS-C and C.HD. An absolute logFC cutoff > 0.1 was employed for differential expression testing.

Finally, to visualize ligand-receptor interactions that are up-regulated in MIS-C, ligand and receptor interactions were plotted where both ligand and receptor connectome logFCs > 1.

### EBV/CMV analysis

To evaluate EBV or CMV infection of individuals in our cohort, we created combined human-viral genome references to align transcriptomic reads and counted the number of detected viral transcripts^39, 40^. To be as permissive as possible, we used CellRanger to map reads to entire viral genomes, to capture counts originating from ORFs, intergenic regions, or initial infection.

### Sub-clustering analysis

Sub-clustering was done on myeloid cells, T and NK cells, and B cells. For T and NK sub-clustering the following clusters were selected: CD4 memory, CD4 naïve I, CD56dim CD16bright NK, CD8 naïve, CD4 naïve II, CD8 memory, gdT cells, MAIT and NKT cells, Regulatory T cells, CD4 and CD8 mixed naive T cells, CD56bright CD16dim NK, Activated memory T cell, Proliferating T and NK cell, NK-T doublets. For myeloid sub-clustering the following clusters were selected: Classical monocytes, Neutrophils, Non-classical monocytes, Platelets, Platelet-T cell doublets, Conventional DC, Platelet-bound monocytes, Plasmacytoid DC, NK-monocyte doublets. For B cell sub-clustering the following clusters were selected: Naïve B, Memory B, Plasma cell, T-NK-B cell doublets. After selecting relevant clusters from the PBMC annotations, we performed the analysis as described above. The same references used to generate PBMC UMAP were applied in the reference-based integration for T and NK cell sub-clustering. For B cell sub-clustering integration, we added an additional reference (C.HD4) due to the unequal donor representation in the activated memory B cell cluster.

For B cell sub-clustering, the first 15 PCs were used for data integration and downstream steps, along with a clustering resolution of 0.3. A.COV5.2 was unable to be integrated into the B cell sub-clustering analysis due to low cell numbers (< 200 B cells) and was removed from this analysis. For T and NK cell sub-clustering, 30 PCs were used for data integration, and 8 PCs for downstream steps, and a clustering resolution of 0.9 was used. For myeloid sub-clustering, 30 PCs were used for data integration, and 15 PCs were used for downstream steps with a clustering resolution of 0.5.

Clusters were annotated as above. Doublet clusters were determined by co-expression of heterogeneous lineage markers (e.g. MS4A1 and CD3D) and nFeature and nCount distribution. Clusters of dead and dying cells were identified as above. Both of these classes of clusters were removed prior to finalizing the UMAPs.

### Cell-type proportion plots

To calculate cell frequencies based on single-cell data, we tabulated donor cells in each cluster, and divided these by the total donor representation in the UMAP. Because of the inherent heterogeneity in our cohorts, a non-parametric two-sided Wilcoxon rank-sum test was used to calculate statistical significance between MIS-C and C.HD.

### DEG analysis and heatmaps

Differentially expressed genes were computed between cohorts using the FindMarkers function in *Seurat*, using the same test and parameters as described above for clusters-specific marker delineation. We used a broader categories of cells to compute differential expression as follows: Monocytes (Classical monocytes I, Classical monocytes II, Classical monocytes III, Intermediate monocytes, Non classical monocytes), Neutrophils (Neutrophils I, Neutrophils II), NK cells (CD56dim S100A4+ NK cells, CD56dim CD38+ NK cells, CD56bright NK cells), CD8 memory (Effector memory CD8 T cells, Central memory CD8 T cells, Terminal effector memory CD8 T cells), Naïve CD4 (Naïve CD4 T cells I, Naïve CD4 T cells II, Naïve CD4 T cells III), Memory CD4 T cells (Memory CD4 T cells, CCR6+ memory CD4 T cells, CXCR3+ memory CD4 T cells),

Naïve B cells (Naïve B, Activated naïve B), Memory B cells (Intermediate memory, Activated memory B, Memory B cell), Plasmablast (Non-dividing plasmablasts, Dividing plasmablasts).

To prioritize genes for analysis, we chose an absolute average log fold-change (logFC) cutoff of an absolute value > 0.5, and a p-adjusted value < 0.05. The top 20 up- or down-regulated genes, sorted by average logFC, were chosen to plot onto heatmaps. Heatmaps were visualized using the *ComplexHeatmap* v2.5.5 package. Correlation heatmaps were created using *Hmisc* v4.4-1 and *corrplot* v0.84.

### Module scores

Module scores were calculated using the AddModuleScore function using the default parameters^41^. The S100 score consists of S100A8, S100A9, and S100A12. The HLA class II score consists of HLA-DRB1, HLA-DRB5, HLA-DRA, HLA-DQA1, HLA-DQA2, and HLA-DQB1.

The super-antigen score includes the following genes: IL2, CXCL9, UBD, IFNG, CXCL11, IL22, ANKRD22, IL17A, IL31RA, FAM26F, CXCL1, IL3, SLAMF8, LOC729936, XCL1, XCL2, SERPING1, SUCNR1, IL27, APOL4, FCGR1A, FCGR1B, SECTM1, CCL8, IL17F, BATF2, GBP4, and ETV7^26^. The viral score consists of SIGLEC1, RABGAP1L, IFI27, CADM1, RSAD2, MX1, and SERPING1^21^. The bacterial score consists of SMPD1, CD44, SERPING1, SPI1, HERC1, MCTP1, FOLR3, CFAP45, PRF1, CTBP1, HLA-DRB1, ARL1, OAS3, ZER1, CHI3L1, IFIT2, and IFITM1^21^.

### Pathway analysis

The MIS-C patients were acutely ill and treated with high-dose steroids, which are known to affect immune gene expression programs. To contend with this noise prior to conducting gene and pathway prioritization, we sought to remove genes that were clearly affected by steroid effects. As a reference, we used a publicly available bulk RNA-seq dataset of PBMC cell-types treated with methylprednisolone, the primary steroid administered to the MIS-C patients^34^. We removed steroid related genes relevant to each cell type from our gene lists, defined as having an absolute logFC cutoff above 2 and p-value less than 0.05 in the steroid dataset and regulated in the same direction as genes in our dataset. Pathway analysis was then done on differentially expressed genes using the above criteria and filtering for steroid-related genes. These differentially expressed genes were inputted to Enrichr^42, 43^ to calculate enrichment of pathway-associated terms.

### BCR analysis

B cell receptor (BCR) repertoire sequence data were analyzed using the Immcantation (www.immcantation.org) framework. Starting with CellRanger output, V(D)J genes for each sequence were aligned to the IMGT reference database v3.1.30^44^ using *IgBlast* v1.13.0^45^. Nonproductive sequences were removed. Within each sample, sequences were grouped into clonal clusters, which contain B cells that relate to each other by somatic hypermutations from a common V(D)J ancestor. Sequences were first grouped by common IGHV gene annotations, IGHJ gene annotations, and junction lengths. Using the DefineClones.py function of *Change-O* v1.0.0^46^, sequences within these groups differing by less than a length normalized Hamming distance of 0.15 within the junction region were defined as clones using single-linkage hierarchical clustering^47^. This threshold was determined by manual inspection of the distance to nearest sequence neighbor distribution for each sample using *Shazam* v1.0.2^19^. These heavy-chain-defined clonal clusters were further split if their constituent cells contained light chains that differed by V and J genes. Within each clone, germline sequences were reconstructed with D segment and N/P regions masked (replaced with “N” nucleotides) using the CreateGermlines.py function within *Change-O* v1.0.0. All BCR analyses used R v3.6.1.

For analysis of B cell clonal diversity, we calculated Simpson’s diversity for each sample using the alphaDiversity function of *Alakazam* v1.0.2^46^. To account for differences in sequence depth, samples within each comparison were down-sampled to the same number of sequences, and the mean of 100 such re-sampling repetitions was reported. Only samples with at least 100 B cells were included.

To identify mutated B cell clones of different cell types and isotypes, B cell clones were further separated by cell type and/or isotype. For all BCR analysis, unless otherwise indicated, “plasmablasts” indicate pooled dividing- and non-dividing annotated plasmablasts defined by sub-clustering annotation and filtered on cells containing BCRs. These B cell clones were considered “mutated” if the median somatic hypermutation frequency of their constituent sequences was ≥ 1%. This threshold is consistent with recent analyses of COVID-19 B cell repertoires^19, 48^.

### TCR analysis

The raw sequencing reads were preprocessed using the Cell Ranger V(D)J pipeline by 10XGenomics™, which assembled read-pairs into V(D)J contigs for each cell, identified cell barcodes from targeted cells, annotated the assembled contigs with V(D)J segment labels and located the CDR3 regions. Only V(D)J contigs with high confidence defined by cell ranger were included for downstream analysis. These V(D)J contigs were re-annotated by aligning them to the IMGT reference database v3.1.3041 using IgBlast v1.13.0 in the Change-O V1.0.0^46^ pipeline. Cells with ambiguous alpha chain or beta chain and cells with no beta chains were removed. For cells with multiple alpha and/or beta chains, if any chain could be captured, we retained the chain with the largest nUMI that provided a unique chain. No sample to sample contamination was identified based on across-cell overlap of cell barcodes and contig sequences.

For analysis of T cell clonal diversity, in each sample, cells with identical alpha chain and beta chain sequences in the repertoire were grouped as one TCR clone. To quantify the clonal diversity, we calculated both Simpson and Shannon diversity for each sample using R package *Alakazam* 1.0.2^46^ which describes the richness and evenness of the repertoire, respectively. When comparing the diversity between samples, to account for the differences in sequencing depth across different samples, down-sampling was conducted to make different samples have the same number of sequences. For each diversity comparison, samples with less than 50 sequences were excluded and all samples were randomly down sampled for 100 times to the smallest number of sequences of the remaining samples. The mean diversity across the 100 times were reported and compared.

We selected memory CD4 and CD8 T cells; naïve CD4 and CD8 T cells and NK cells for downstream analysis. We separated Ki67+ cells into CD4 and CD8 categories and combined these with the memory CD4 and CD8 cells, respectively.

The principle component analysis (PCA) was performed on TRBV gene frequency within each individual using R package *stats* v4.0.2. Statistical significance of PCA clustering is determined by permutation test, where the statistic is the ratio of mean intra/inter cluster Euclidean distances among points. Mean values of the fraction of each TRBV gene per sample with error bars are used to demonstrate the differential gene usage. TRBV genes are sorted according on the difference between MIS-C-S and C.HD in descending order.

### Serum antibody binding to cultured endothelial cells

De-identified and discarded high-titer panel reactive antigen (PRA) sera showing >80% reactivity to HLA Class-I and II antigens were collected from transplant patients at Yale New Haven Hospital’s tissue typing laboratory. Healthy donor (HD) and moderate and severe MIS-C patient serum was isolated by centrifugation of serum collection tubes at 840g for 10 minutes. Supernatant was flash frozen in liquid nitrogen and stored in -80C prior to being thawed for experiments. To induce *in situ* levels of HLA antigens expression, 60% confluent human cardiac microvascular endothelial cells (HCMECs, Lonza) were pre-treated with human recombinant IFN-γ (final concentration of 100 units/mL, 48 hours, Invitrogen) and TNF! (final concentration of 10 ng/mL for 6-8 hours, Invitrogen) in EGM2 MV Microvascular Endothelial Cell Growth Medium-2 (Lonza CC-4147). Cells were then washed with HBSS (Gibco) and treated with PRA, HD or MIS-C serum in a 1:1 ratio with gelatin veronal buffer (GVB, Sigma) for 2 hours. Untreated cells were washed with HBSS (Gibco) followed by addition of GVB for 2 hours. To assess antibody binding, cells were suspended in trypsin, washed in 1% BSA PBS (FACS buffer) and pelleted by centrifugation at 1000 rpm. The cells were then incubated with goat anti-human IgG (H+L) cross-adsorbed secondary antibody, alexa fluor 594 (ThermoFisher, Catalog # A-11014) at 1:400 dilution with FACS buffer. Unbound antibodies were removed by washing three times with 1xPBS (Gibco) followed by fixing with 3.7% Formaldehyde solution (T Baker catalogue# 2106-01) at room temperature. Cells were washed twice in FACS buffer and resuspended in 200-300 uL FACS buffer for flow cytometric analysis.

### Serum protein analysis

Relative serum protein levels were quantified by the SOMAscan v4 platform (Somalogic Inc., Boulder CO), which measures the binding of 5,284 modified single-stranded DNA aptamers (SOMAmers) to specific analytes in each sample (Gold et al., 2010). The assay and characteristics of the reagents have been described before (Emilsson et al., 2018). Briefly, 120µl aliquots of serum samples from three MIS-C, one Kawasaki, and one toxic shock syndrome (TSS) patients were placed across two 96-well plates with four age- and gender-matched C.HD, independent controls, and other samples not analyzed in this study. C.HD used in serum analysis were distinct from C.HD used in scRNAseq analysis. All samples were sent together on dry ice and assayed by Somalogic. Measurements were standardized using the default method performed by the manufacturer to first account for hybridization and assay bias within plates, followed by plate scaling and calibration to remove plate effects, and median normalization to a reference at the end. Based on a subset of 4,706 human protein analytes (representing 4,478 distinct proteins) that passed quality control, the median intra-subject coefficient of variation (CV) of 3.30% from eight blinded technical replicates across two plates suggests low technical variability.

Serum connectivity networks were created by selecting the top 40 differentially up-regulated proteins in MIS-C compared to C.HD, and mapping them to receptors expressed in PBMC using the ligand-receptor reference used previously. We defined a receptor as being expressed in PBMCs if it is expressed in at least 25% of cells in any cluster.

### Statistical tests

The number of samples per group and experiment repeats, as well as the statistical test used is indicated in each figure legend.

## SUPPLEMENTAL INFORMATION

**Table S1.** Patient characteristics. * asthma, seizures, developmental delay, substance abuse/mental illness, 1 critical patient with chronic kidney disease (CKD) and chronic heart failure (CHF) diagnosed at the time of MIS-C work up. ** 1 patient IgG not checked *** 1 patient with group A strep found on throat culture + Length of Stay for one patient excluded as still admitted

| **Characteristic** | **All MIS-C**<br>**(n=15)** | **Severe**<br>**(n=8)** | **Moderate**<br>**(n=7)** |
| --- | --- | --- | --- |
| **Age (years)** | 10.4 (2.6-18) | 12 (3-18) | 8.7 (2.6-17) |
| **Sex: Male: Female** | 8:7 | 5:3 | 3:4 |
| **Race: Black** | 3 (20%) | 1 (13%) | 2 (29%) |
| **Hispanic/**<br>**Latino** | 12 (80%) | 7 (88%) | 5 (71%) |
| **White** | 0 (0%) | 0 (0%) | 0 (0%) |
| **Body Mass Index (kg/m<sup>2</sup>)** | 23.3 (13.8-31.2) | 23.7 (18.6-31.2) | 22.9 (13.8-30.3) |
| **Past Medical History\*** | 6 (40%) | 2 (25%) | 4 (57%) |
| **Known COVID+ contact** | 5 (33%) | 1 (13%) | 4 (57%) |
| **SARS/CoV2 PCR+** | 6 (40%) | 4 (50%) | 2 (29%) |
| **SARS/CoV2 IgG+** | 14 (100%) | 7 (100%)** | 7 (100%) |
| **Other infection\*\*\*** | 1 (7%) | 1 (13%) | 0 (0%) |
| **Clinical features** |  |  |  |
| **Fever** | 14 (93.3%) | 7 (88%) | 7 (100%) |
| **GI:** |  |  |  |
| **Abdominal pain** | 11 (73%) | 5 (63%) | 6 (86%) |
| **Emesis** | 13 (87%) | 7 (88%) | 6 (86%) |
| **Diarrhea** | 12 (80%) | 8 (100%) | 4 (57%) |
| **Cardiovascular:** |  |  |  |
| **Chest pain** | 3 (20%) | 2 (25%) | 1 (14%) |
| **Cardiogenic Shock** | 6 (40%) | 6 (75%) | 0 (0%) |
| **Distributive Shock** | 7 (46.6%) | 7 (88%) | 0 (0%) |
| **Neurologic:** |  |  |  |
| **Headache** | 6 (40%) | 3 (38%) | 3 (43%) |
| **Confusion** | 3 (20%) | 0 (0%) | 3 (43%) |
| **Rash** | 10 (67%) | 6 (75%) | 4 (57%) |
| **Conjunctivitis** | 9 (60%) | 6 (75%) | 3 (43%) |
| **Sore throat** | 4 (27%) | 2 (25%) | 2 (29%) |
| **Muscle aches** | 4 (27%) | 3 (38%) | 1 (14%) |
| **Lymphadenopathy** | 0 (0%) | 0 (0%) | 0 (0%) |
| **Diagnostics** |  |  |  |
| **ECHO:** |  |  |  |
| **Depressed Left Ventricular**<br>**function** | 6 (40%) | 5 (63%) | 1 (14%) |
| **Coronary aneurism**<br>**(z score>2)** | 2 (13%) | 2 (25%) | 0 (0%) |
| **Therapy** |  |  |  |
| **Respiratory Support:** |  |  |  |
| **Intubation** | 2 (13%) | 2 (25%) | 0 (0%) |
| **Non-invasive PPV** | 1 (7%) | 1 (13%) | 0 (0%) |
| **Oxygen support** |  |  |  |
| **Regular NC** | 1 (7%) | 0 (0%) | 1 (14%) |
| **HFNC** | 2 (13%) | 2 (25%) | 0 (0%) |
| **Corticosteroids** | 15 (100%) | 8 (100%) | 7 (100%) |
| **Intravenous Immunoglobulin** | 13 (87%) | 8 (100%) | 5 (71%) |
| **Anakinra** | 7 (47%) | 5 (63%) | 2 (29%) |
| **Tocilizumab** | 1 (7%) | 1 (13%) | 0 (0%) |
| **Remdesivir** | 1 (7%) | 1 (13%) | 0 (0%) |
| **Heparin** | 7 (46%) | 5 (63%) | 2 (29%) |
| **Aspirin** | 15 (100%) | 8 (100%) | 7 (100%) |
| **Antibiotics** | 11 (73%) | 7 (88%) | 4 (57%) |
| **Convalescent plasma** | 1 (7%) | 1 (13%) | 0 (0%) |
| **Vasoactive medication** | 7 (47%) | 7 (88%) | 0 (0%) |
| **Outcomes** |  |  |  |
| **Length of Stay, days** | 6.9 (2-15)<sup>+</sup><br>n=14 | 8.5 (6-15)<sup>+</sup><br>n=7 | 5.4 (2-9) |
| **Death** | 0 | 0 | 0 |

**Table S2.** Clinical laboratory tests. BNP-B-type natriuretic peptide; AST-aspartate aminotransferase; ALT-alanine aminotransferase; CRP-c-reactive protein. * 1 patient CD25 likely too high to measure

| Characteristic | All MIS-C<br>(n=15) | Severe<br>(n=8) | Moderate<br>(n=7) |
| --- | --- | --- | --- |
| **Laboratory Tests:** |  |  |  |
| **Ferritin, ng/mL** | 2,035 (100-12,823) | 3,278 (138-12,823) | 614 (100-2,615) |
| **BNP, pg/mL** | 12,568 (2,065->70,000)<br>n=14 | 21,313 (2,653->70,000)<br>n=7 | 3,823 (2,065-5,752) |
| **Troponin, ng/mL** | 0.10 (<0.01-0.40)<br>n=14 | 0.15 (0.015-0.40) | 0.02 (<0.01-0.06)<br>n=6 |
| **Creatinine, mg/dL** | 1.80 (0.31-11.17) | 2.36 (0.40-11.17) | 1.16 (0.31-4.70) |
| **AST, U/L** | 291 (23-2,799) | 395 (23-2,799) | 173 (29-552) |
| **ALT, U/L** | 169 (14-1,343) | 225 (28-1,343) | 107 (14-280) |
| **Albumin, g/dL** | 2.6 (2.0-3.7) | 2.3 (2.0-2.8) | 3.0 (2.4-3.7) |
| **Lactate, mmol/L** | 4.1 (0.9-14.0)<br>n=12 | 5.2 (1.1-14.0) | 1.9 (0.9-3.8)<br>n=4 |
| **D-dimer, mg/L** | 5.1 (1.5-21.0) | 7.0 (2.4-21.0) | 3.1 (1.5-6.6) |
| **CRP, mg/L** | 187 (6->300) | 178 (6->300) | 197 (93->300) |
| **Absolute Lymphocyte Count, x1000/uL** | 0.7 (0.0-1.8) | 0.6 (0.0-1.8) | 0.8 (0.3-1.3) |
| **Procalcitonin, ng/mL** | 12.0 (0.1-36.6)<br>n=14 | 11.4 (0.1-31.3)<br>n=7 | 12.6 (1.5-36.6) |
| **CD25, pg/mL** | 15,723 (1,565-48,300)<br>n=4 | 20,442 (2,598-48,300)<br>n=3* | 1,565<br>n=1 |
| **IL-6, pg/mL** | 45.9 (<2.0-147.0)<br>n=9 | 58.5 (5.0-147.0)<br>n=6 | 20.8 (1.8-58.6)<br>n=3 |
| **IL-10, pg/mL** | 126.5 (7.4-259.0)<br>n=5 | 137.9 (7.4-259.0)<br>n=4 | 80.9<br>n=1 |
| **IFNγ, pg/mL** | 6.3 (<4.2-13.0)<br>n=5 | 6.8 (<4.2-13.0)<br>n=4 | <4.2<br>n=1 |

**Table S3.** Characteristics of patients included in single-cell RNA sequencing. M-male; F-female; DD-developmental delay; CKD-chronic kidney disease; CHF-chronic heart failure; Hisp-Hispanic; Methylpred-methylprednisolone; IVIG-intravenous immunoglobulin.

| ID | BMI (kg/m<sup>2</sup>) | Medical History | Other Infection | Symptoms | Immune modulation prior to blood sample |
| --- | --- | --- | --- | --- | --- |
| P1 | 19.4 | None | Throat: Group A Strep | Fever, rash, conjunctivitis, abdominal pain, vomiting, diarrhea, myalgia, sore throat | Methylpred, IVIG, anakinra |
| P2 | 22 | Smoker | None | Fever, rash, conjunctivitis, vomiting, diarrhea, shortness of breath | Methylpred, IVIG, aspirin, convalescent plasma, anakinra |
| P3 | 24.6 | None | None | Fever, rash, conjunctivitis, abdominal pain, vomiting, diarrhea, chest pain | Remdesivir, anakinra |
| P4 | 21.4 | None | None | Fever, rash, abdominal pain, vomiting, diarrhea | Methylpred, aspirin |
| P5 | 17.2 | Seizures | None | Fever, abdominal pain, vomiting, diarrhea | Methylpred, aspirin |
| P6 | 29.8 | DD, CKD, CHF | None | Chills, shortness of breath, cough, abdominal pain, vomiting, diarrhea, anorexia, myalgia, sore throat, headache | Methylpred, anakinra |
| P7 | 32.3 | None | None | Fever, rash, fatigue, vomiting, diarrhea, headache and altered mental status | Methylpred, IVIG, aspirin, anakinra |

**Table S4.** Meta-data for sequencing samples. Indicated are unique sample IDs, subject IDs, patient characteristics (age, sex, condition, icu status etc.), and processing conditions. Also indicated are the lab from which the samples originated (where Lucas1 and Lucas2 batches were processed on different days), the number of days between hospitalization and blood draw (no_days_bt_hosp_blood_draw), and the number of hours between blood draw and processing by Ficoll (no_hrs_bt_draw_and_ficoll).

| Sample ID | Condition | Subject ID | Time point | # days between hosp. and draw | Sex | Processing Lab | Storage | Blood shipped o/n (y/n) | # hours between draw and ficoll | Age group | Donor/Patient | ICU status | Severity |
| --- | --- | --- | --- | --- | --- | --- | --- | --- | --- | --- | --- | --- | --- |
| P1.1 | MIS-C | P1 | A | 4 | M | Lucas1 | fresh | N | less than 3 | Ped | Pt | Y | MIS-C-S |
| TSS | TSS | TSS | A | 1 | F | Lucas1 | fresh | N | less than 3 | Ped | Pt | Y | NA |
| P2.1 | MIS-C | P2 | A | 4 | M | Lucas1 | fresh | N | less than 3 | Ped | Pt | Y | MIS-C-S |
| A.HD1 | A.HD | A.HD1 | NA | NA | M | Lucas1 | fresh | N | less than 3 | Adult | HD | NA | NA |
| A.HD2 | A.HD | A.HD2 | NA | NA | F | Lucas1 | fresh | N | less than 3 | Adult | HD | NA | NA |
| A.HD3 | A.HD | A.HD3 | NA | NA | M | Lucas1 | fresh | N | less than 3 | Adult | HD | NA | NA |
| A.COV1.1 | COVID19-A | A.COV1 | A | 2 | M | Kaminski | cryopreserved | N | NA | Adult | Pt | N | NA |
| A.COV1.2 | COVID19-B | A.COV1 | B | 6 | M | Kaminski | cryopreserved | N | NA | Adult | Pt | N | NA |
| A.COV2.1 | COVID19-A | A.COV2 | A | 3 | F | Kaminski | cryopreserved | N | NA | Adult | Pt | N | NA |
| A.COV2.2 | COVID19-B | A.COV2 | B | 6 | F | Kaminski | cryopreserved | N | NA | Adult | Pt | N | NA |
| A.COV3.1 | COVID19-A | A.COV3 | A | 3 | M | Kaminski | cryopreserved | N | NA | Adult | Pt | N | NA |
| A.COV3.2 | COVID19-B | A.COV3 | B | 15 | M | Kaminski | cryopreserved | N | NA | Adult | Pt | N | NA |
| A.COV4.1 | COVID19-A | A.COV4 | A | 2 | F | Kaminski | cryopreserved | N | NA | Adult | Pt | N | NA |
| A.COV4.2 | COVID19-B | A.COV4 | B | 6 | F | Kaminski | cryopreserved | N | NA | Adult | Pt | N | NA |
| A.COV5.2 | COVID19-B | A.COV5 | B | 12 | M | Kaminski | cryopreserved | N | NA | Adult | Pt | Y | NA |
| A.COV6.2 | COVID19-B | A.COV6 | B | 8 | M | Kaminski | cryopreserved | N | NA | Adult | Pt | Y | NA |
| A.HD4 | A.HD | A.HD4 | NA | NA | M | Kaminski | cryopreserved | N | NA | Adult | HD | NA | NA |
| A.HD5 | A.HD | A.HD5 | NA | NA | M | Kaminski | cryopreserved | N | NA | Adult | HD | NA | NA |
| A.HD6 | A.HD | A.HD6 | NA | NA | M | Kaminski | cryopreserved | N | NA | Adult | HD | NA | NA |
| A.HD7 | A.HD | A.HD7 | NA | NA | M | Kaminski | cryopreserved | N | NA | Adult | HD | NA | NA |
| A.HD8 | A.HD | A.HD8 | NA | NA | F | Hafler | fresh | N | NA | Adult | HD | NA | NA |
| A.HD9 | A.HD | A.HD9 | NA | NA | F | Hafler | fresh | N | NA | Adult | HD | NA | NA |
| A.HD10 | A.HD | A.HD10 | NA | NA | F | Hafler | fresh | N | NA | Adult | HD | NA | NA |
| A.HD11 | A.HD | A.HD11 | NA | NA | M | Hafler | fresh | N | NA | Adult | HD | NA | NA |
| A.HD12 | A.HD | A.HD12 | NA | NA | F | Hafler | fresh | N | NA | Adult | HD | NA | NA |
| A.HD13 | A.HD | A.HD13 | NA | NA | M | Hafler | fresh | N | NA | Adult | HD | NA | NA |
| P3.1 | MIS-C | P3 | A | 2 | M | Lucas2 | cryopreserved | N | less than 6 | Ped | Pt | Y | MIS-C-S |
| P4.1 | MIS-C | P4 | A | 2 | F | Lucas2 | cryopreserved | N | less than 6 | Ped | Pt | N | MIS-C-M |
| P5.1 | MIS-C | P5 | A | 2 | F | Lucas2 | cryopreserved | N | less than 6 | Ped | Pt | N | MIS-C-M |
| P6.1 | MIS-C | P6 | A | 3 | M | Lucas2 | cryopreserved | N | less than 6 | Ped | Pt | Y | NA |
| P7.1 | MIS-C | P7 | A | 1 | M | Lucas2 | cryopreserved | Y | about 24 | Ped | Pt | Y | MIS-C-S |
| P3.2 | MIS-C-R | P3 | B | 73 | M | Lucas2 | cryopreserved | N | less than 6 | Ped | Pt | NA | NA |
| P4.2 | MIS-C-R | P4 | B | 45 | F | Lucas2 | cryopreserved | N | less than 6 | Ped | Pt | NA | NA |
| C.HD1 | C.HD | C.HD1 | NA | NA | F | Lucas2 | cryopreserved | N | less than 6 | Ped | HD | NA | NA |
| C.HD2 | C.HD | C.HD2 | NA | NA | M | Lucas2 | cryopreserved | Y | about 24 | Ped | HD | NA | NA |
| C.HD3 | C.HD | C.HD3 | NA | NA | F | Lucas2 | cryopreserved | N | less than 6 | Ped | HD | NA | NA |
| C.HD4 | C.HD | C.HD4 | NA | NA | M | Lucas2 | cryopreserved | Y | about 24 | Ped | HD | NA | NA |
| C.HD5 | C.HD | C.HD5 | NA | NA | M | Lucas2 | cryopreserved | Y | about 24 | Ped | HD | NA | NA |
| C.HD6 | C.HD | C.HD6 | NA | NA | M | Lucas2 | cryopreserved | Y | about 24 | Ped | HD | NA | NA |

Table S5. CITE-seq panel.

189 markers used for CITE-seq. Listed are the surface markers, corresponding gene names, and associated sequences.

Table S6. Differentially expressed genes.

Differential expression markers between MIS-C and C.HD, in clusters corresponding to broad immune cell subsets. (see methods). Markers gated on abs(logFC) > 0.05 and p.adj < 0.05.

**Figure S1.**
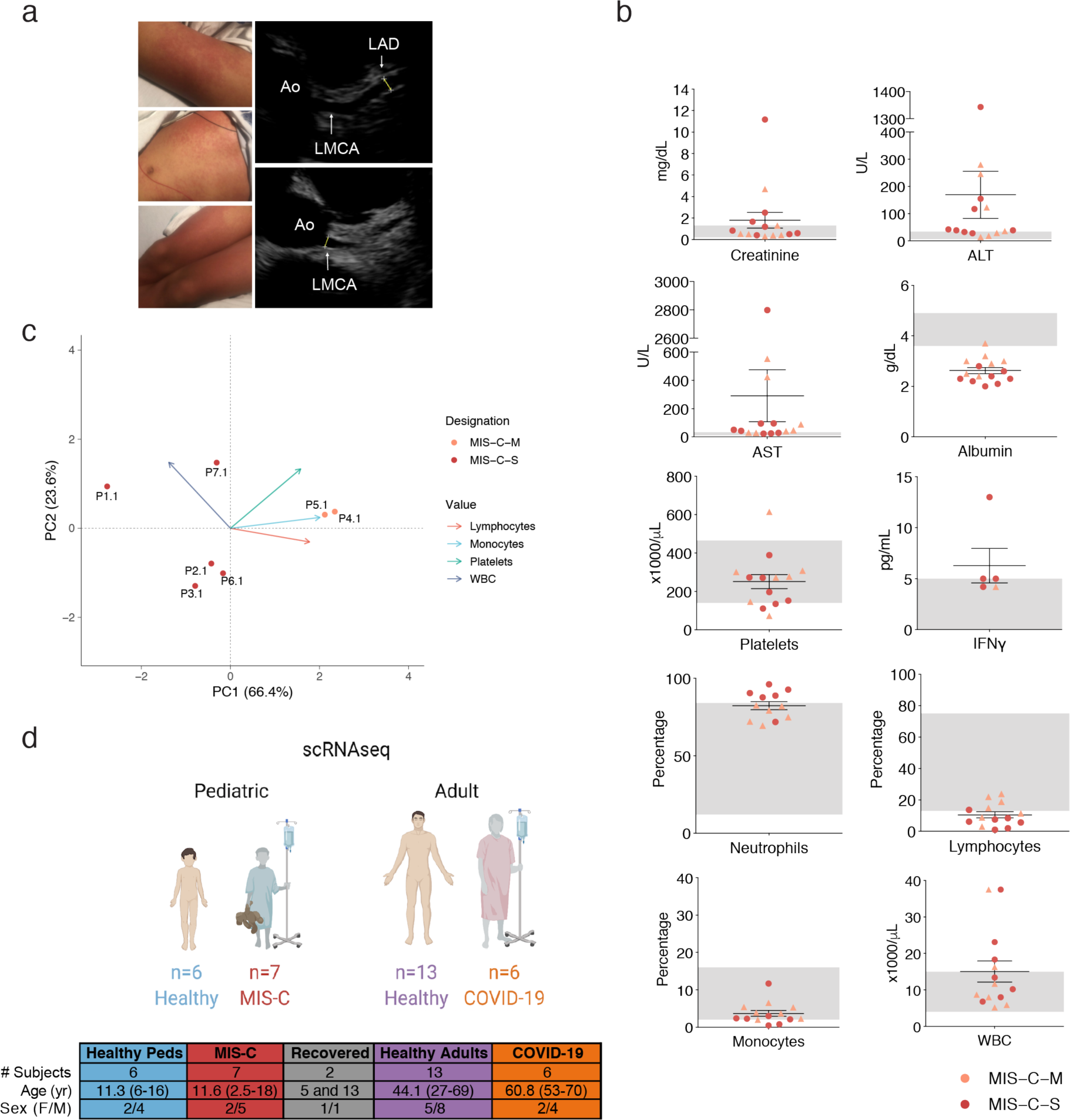
Supplemental clinical features. (**a**) MIS-C rash on patient P1 and coronary aneurism in left anterior descending coronary artery, Z score of 3.51, in patient P3. Ao-aorta; LMCA-left main coronary artery; LAD-left anterior descending coronary artery. (**b**) Acute phase laboratory values (creatinine, ALT/AST, albumin, IFN!) and laboratory values on the day of blood sampling/protocol consent (WBC, neutrophils, lymphocytes, monocytes). Severe MIS-C (MIS-C-S) is highlighted in red dots and moderate MIS-C (MIS-C-M) in light red triangles. Normal range represented by gray shading. AST-aspartate aminotransferase; ALT-alanine aminotransferase; WBC-white blood cells. (**c**) PCA plot of single-cell RNA sequencing cohort separates severe and moderate patients by complete blood count values at time of blood sampling. (**d**) Overview of single-cell RNA sequencing cohorts.

**Figure S2.**
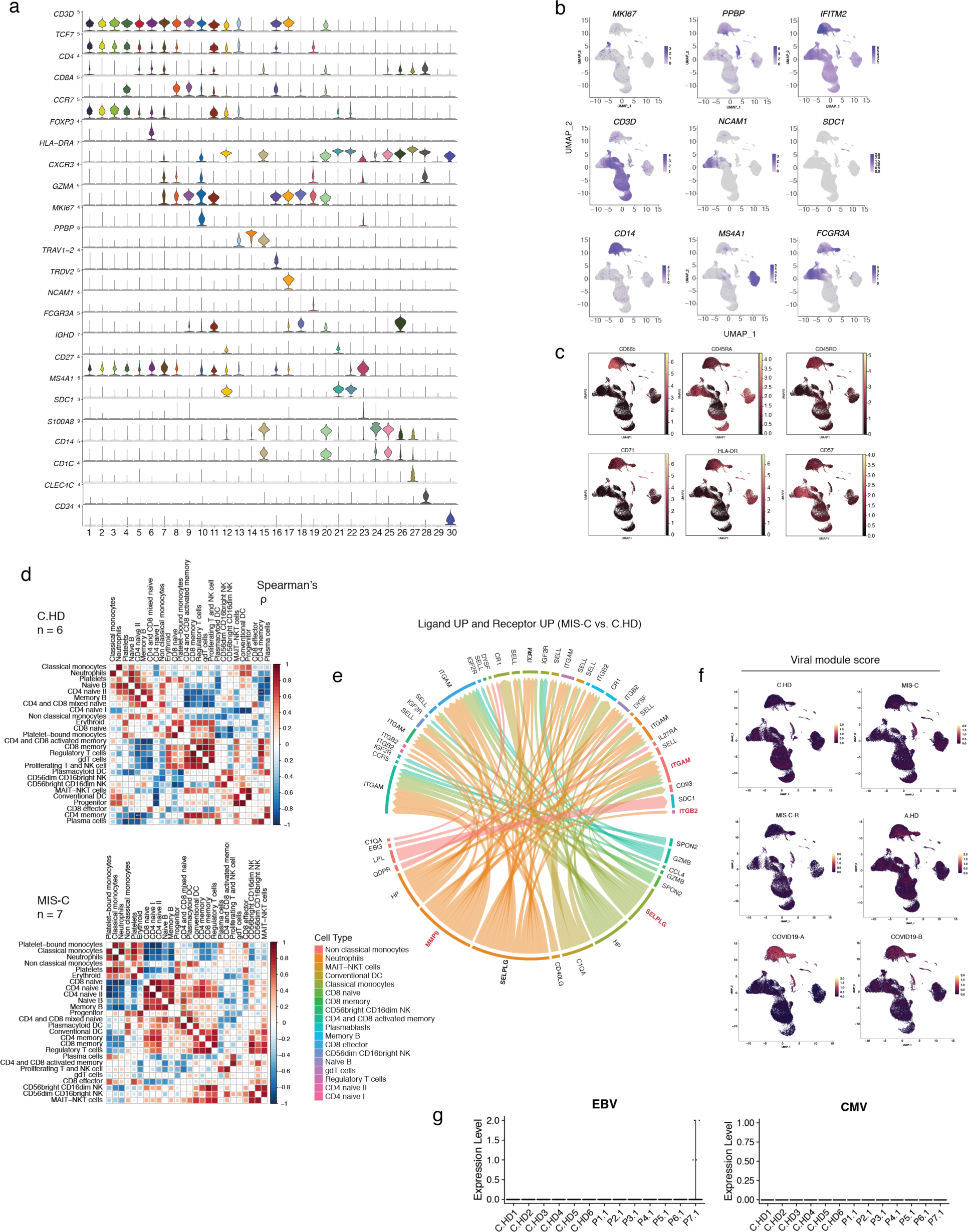
Comprehensive analysis of PBMC clusters, receptor-ligand pairs, and viral gene modules. (**a**) Violin plots depicting extensive PBMC cell lineage markers, as in Figure 2b. (**b**) UMAP overlay of markers delineating major cell types including T cells, NK cells, plasma cells, B cells, monocytes, and neutrophils. Scale represents normalized GEX feature counts. (**c**) PBMC UMAPs with overlay of CITE-seq data. Scale represents normalized CITE-seq feature counts. (**d**) Correlation matrix of cell frequencies within C.HD (top) and within MIS-C cohorts (bottom). Scale represents Spearman’s rho. P-values, where depicted, were calculated using Wilcoxon rank sum test, and adjusted for multiple comparisons using the Benjamini Hochberg procedure^49^. *** indicates p < 0.001. (**e**) Predicted ligand-receptor interactions from genes that are significantly up-regulated in MIS-C vs. C.HD. Highlighted are ligands and receptors involved in diapedesis and/or inflammation. (**f**) Viral and bacterial gene expression scores in PBMC UMAP, split by condition. (**g**) Counts mapping to EBV and CMV reference transcriptomes.

**Figure S3.**
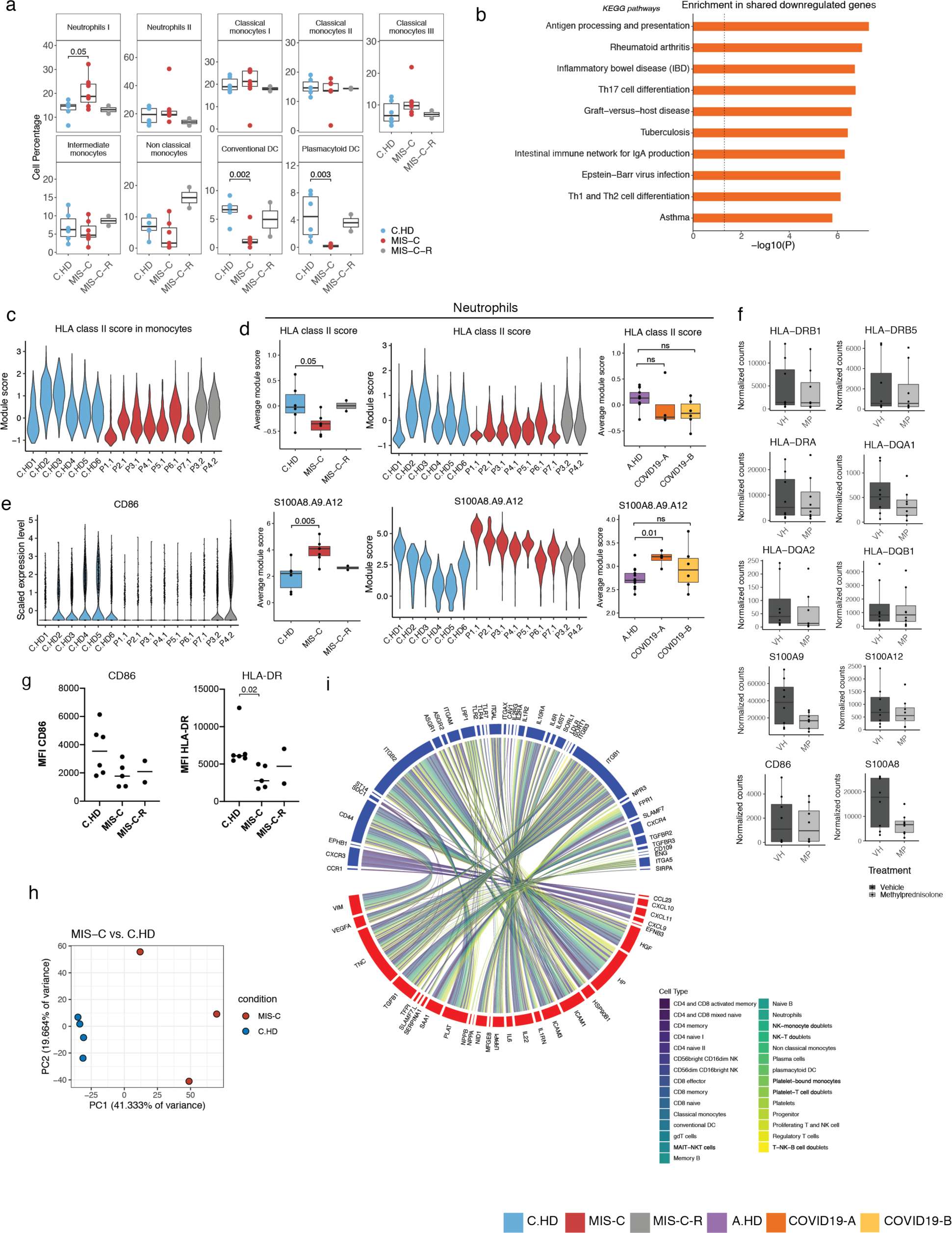
Supplemental myeloid cell findings and serum ligand-PBMC receptor connectome. (**a**) Myeloid cell type percentages across donors. (**b**) Pathways enriched in down-regulated differentially expressed genes shared by monocytes and neutrophils between MIS-C vs. C.HD. (**c**) HLA class II score across donors in monocytes (**d**) HLA class II score (top) and *S100A8*, *A9*, and *A12* (bottom) in neutrophils across pediatric and adult donors. (**e**) *CD86* across donors in myeloid cells. (**f**) *S100*-, *CD86*, and *HLA* gene expression changes are quantified based on a publicly available RNA-sequencing data of *in vitro* steroid treatment of myeloid cells for 6 hours with methylprednisolone or DMSO^34^. (**g**) CD86 and HLA-DR flow cytometry data of pediatric donor samples, gated on monocytes. Statistical significance computed with a two-tailed t-test. (**h**) PCA of individuals based on serum proteomic data. Conditions healthy pediatric donors (n = 4) and MIS-C patients (n = 3). (**i**) Connectivity network representing top 40 differentially expressed serum ligands and receptor pairs, where receptors are expressed in at least one PBMC cluster (minimum percentage cutoff = 0.25). Ribbon colors represent receptor-associated cell type.

**Figure S4.**
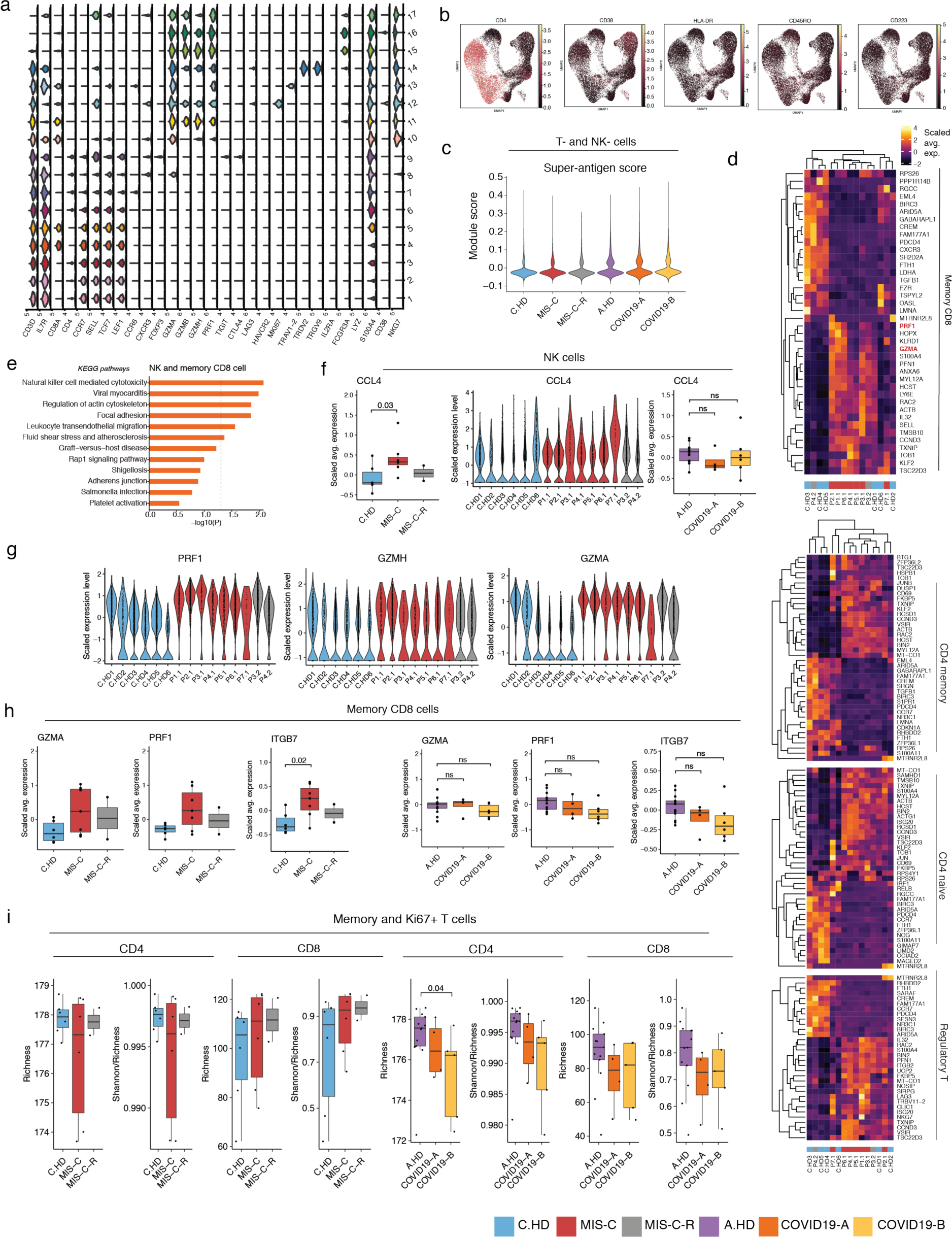
Supplemental NK and T cell findings. (**a**) Violin plots depicting extensive T cell sub-clustering lineage markers. (**b**) T cell UMAPs with overlay of CITE-seq data. (**c**) Super-antigen module score depicted across T and NK cells. (**d**) Heatmap representing top differential expressed genes between MIS-C vs C.HD in memory CD8 T cells (top) and CD4 T cell subsets (bottom). (**e**) Analysis of pathways using Enrichr for shared up-regulated genes in NK and memory CD8 T cells. (**f**) *CCL4* expression in NK cells across pediatric and adult donors. (**g**) *PRF1*, *GZMA*, and *GZMH* expression in NK cells across pediatric donors. (**h**) *GZMA*, *PRF1*, and *ITGB7* in memory CD8 cells across pediatric and adult donors. (**i**) Rarefied diversity indices (richness and evenness) of combined Ki67+ and memory T cells in TCR data analysis.

**Figure S5.**
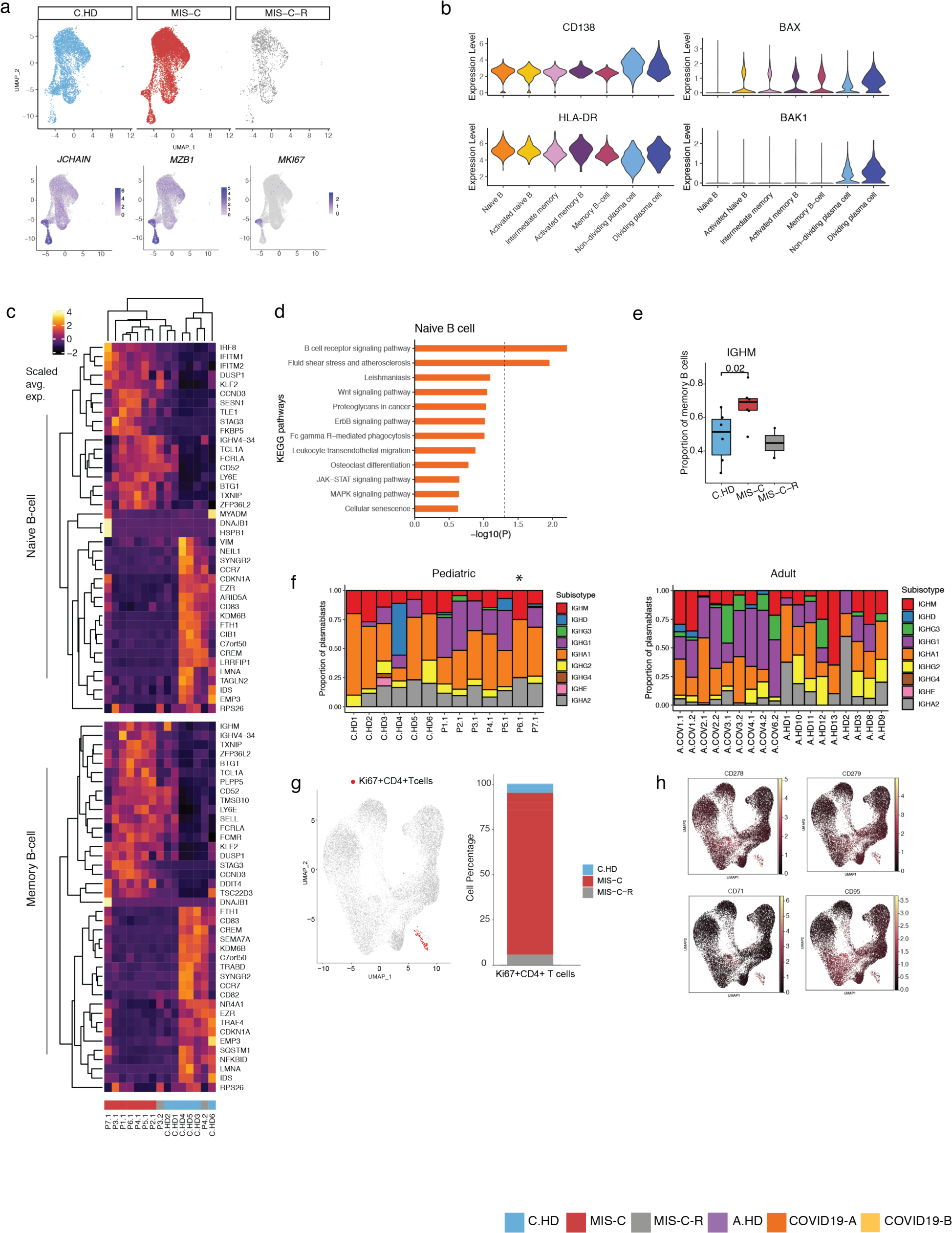
Supplemental B cell findings. (**a**) B cell UMAP split across pediatric conditions (C.HD, MIS-C, and MIS-C-R) (top). Marker genes delineating dividing plasmablasts are overlayed onto fully integrated UMAP (bottom). (**b**) Markers used to annotate short-lived plasmablasts among B cell clusters. (**c**) Heatmap depicting differential expressed genes in naïve B cells and memory B cells. (**d**) Pathway analysis of up-regulated differentially expressed genes in naïve B cells. (**e**) Proportion of IGHM+ memory B cells by analysis of constant regions (**f**) Isotype compositions of pediatric and adult cohorts. *P6.1 also had chronic kidney and heart disease. (**g**) Ki67+ CD4 T cells are labeled on T cell UMAP (left). Proportion of Ki67+ CD4 T cells across pediatric cohorts (right). (**h**) CITE-seq overlay on T cell UMAP depicting expression of B helper surface markers in the Ki67+ CD4 T cells.

**Figure S6.**
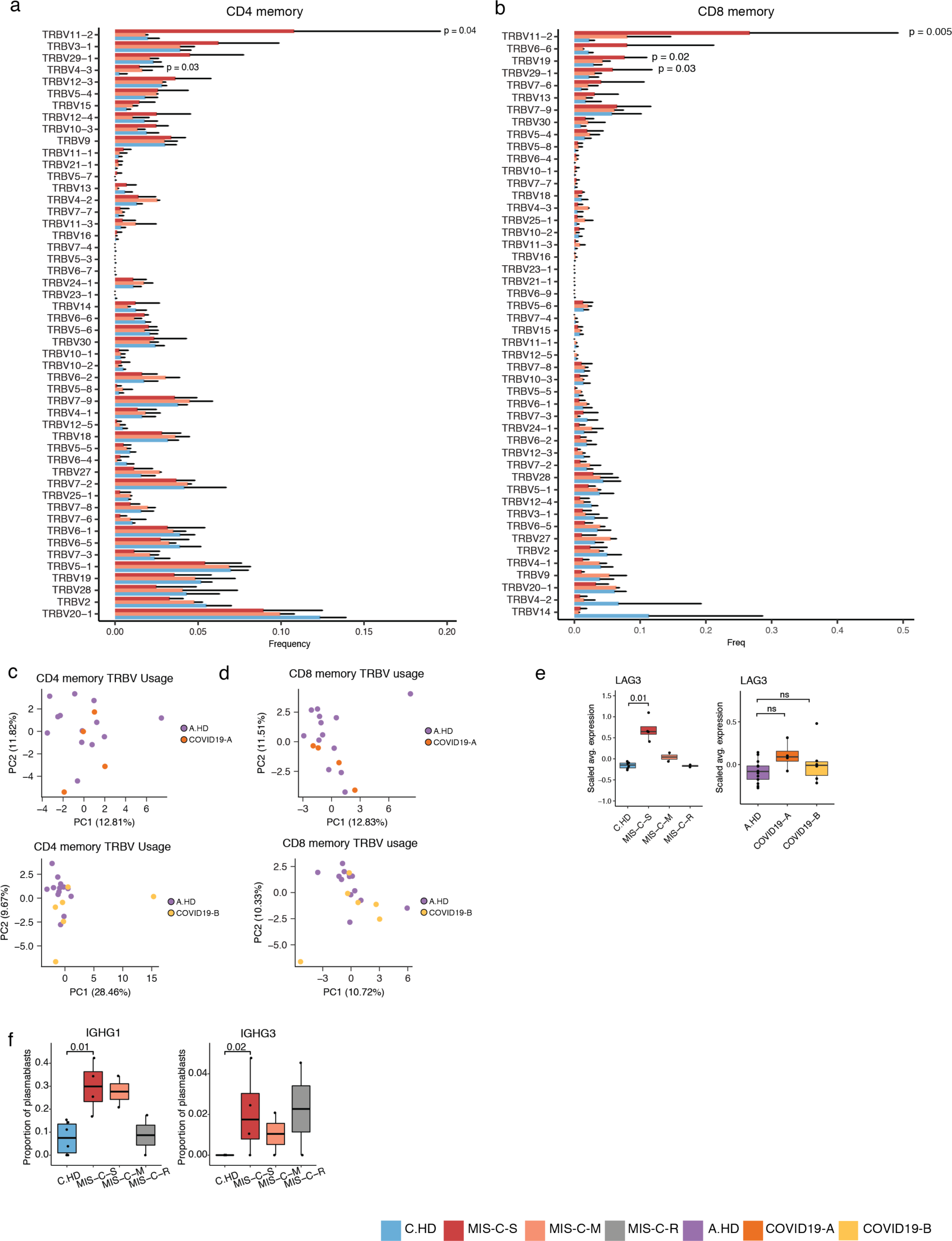
Supplemental findings in severe MIS-C patients. (**a**) Distribution of *TRBV* gene usage in CD4 memory (see **Figure 6a**) with statistical significance computed between MIS-C-S and C.HD using a two-sided Wilcoxon rank sum test. (**b**) As in (a), for CD8 memory. (**c**) PCA of *TRBV* usage in CD4 memory cells for A.HD and COVID19-A (top) and A.HD and COVID19-B (bottom). (**d**) As in (c), for CD8 memory. (**e**) *LAG3* expression across pediatric and adult donors. (**e**) Proportion of *IGHG1* and *IGHG3* plasmablasts compared between C.HD and MIS-C-S.

**Figure S7.**
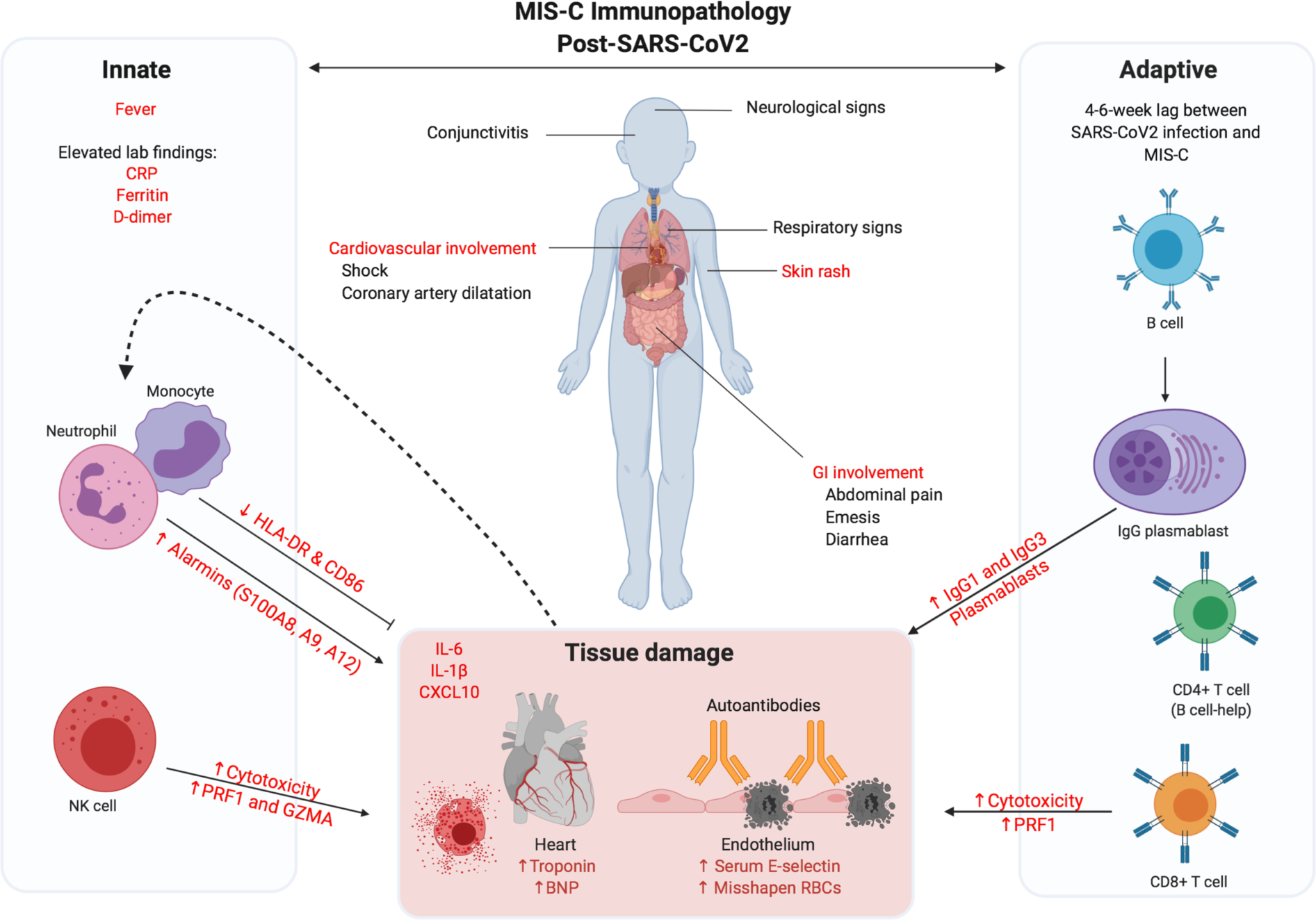
Schematic model of immunopathology drivers in MIS-C. Figure generated with Biorender.com.

